# Parental acceptance toward behavior guidance techniques for pediatric dental visits: a meta-analysis

**DOI:** 10.1101/2020.10.13.20212191

**Authors:** Carla Massignan, Josiane Pezzini Soares, Maria Marlene de Souza Pires, Bruce D. Dick, André Luís Porporatti, Graziela De Luca Canto, Michele Bolan

## Abstract

**Objective:** The systematic review aimed to compare agreement with behavior guidance techniques (BGT) between parents of children with special health care needs (SHCN) and those non-SHCN.

**Methods:** A structured search of Cochrane Library, Latin American and Caribbean Health Sciences, PubMed, PsycInfo, Scopus, Web of Science, ProQuest Dissertations and Theses Database, Opengrey and Google Scholar was taken up to October 2020. Two authors selected studies independently, extracted the data, assessed the studies’ methodological quality using the Joanna Briggs scale and the Recommendations, Assessment, Development and Evaluation (GRADE).

**Results:** Forty-eight studies covering the parents’ agreement with BGT were included and 41 were retained for random-effects proportion meta-analysis. The methodological quality assessment varied from low to high. Among the parents of non-SHCN children, the agreement with BGT varied from 84.1% (95% CI: 75.8-90.9; p<0.001; I^2^ 93.3%) for tell-show-do to 25.7% (95% CI: 17.8-34.4; p<0.001; I^2^ 90.4%) for passive protective stabilization, without hand-over-mouth. Among the parents of children with SHCN, the acceptance of BGT varied from 89.1% (95% CI: 56.1-99.7; p<0.001; I^2^ 95.7%) for tell-show-do to 29.1% (95% CI: 11.8-50.0; p=0.001; I^2^ 84.8%) to general anesthesia.

**Conclusion:** There is very low certainty in evidence that both the parents of children SHCN and non-SHCN were more likely to agree with basic BGT and that they were less likely to agree with the advanced ones.

## Introduction

The long-term success of any dental treatment provided for children depends on the behavior guidance technique (BGT). The dentist approach needs to be integrated to the overall BGT use, taking into account children’s individuality, the practitioner’s skills and the parents’ opinion^2^. Given the changes in the society in the past years where more fathers, mothers, and siblings are accompanying children to their dental appointments^3^, there is considerable interest of families to take part of the treatment decisions. As a result, the attitudes of modern parents have influenced the use of BGT^4^.

The techniques utilized by the dental team have evolved along the years accompanying the society and parenting changes^4^. Currently, according to American Academy of Pediatric Dentistry (AAPD), the BGT are divided in basic behavior guidance, which include communication and communicative guidance; positive pre-visit imagery; direct observation; tell-show-do; ask-tell-ask; voice control; nonverbal communication; positive reinforcement and descriptive praise; distraction; memory restructuring; parental presence/absence; communication techniques for parents and age appropriate patients; and nitrous oxide/oxygen inhalation; and advanced behavior guidance which include protective stabilization, sedation, and general anesthesia^6^. Furthermore, protective stabilization can involve another person, a device or a combination thereof^5^.

Behavioral guidance techniques are used to reduce anxiety and fear, establish a positive attitude, and provide oral health care with physical and emotional security for children with and without special health care needs children (SHCN)^5^. Some patients find it very difficult to cooperate during treatment and the use of only non-pharmacological techniques may be insufficient. In such cases, behavior guidance can be individualized according to the patient’s needs and the parents’ preferences^6^.

Considering that the treatment plan also depends on the parents’ opinion about BGT use, exploring parents’ opinions is critical when identifying BGT application priorities. More invasive procedures can produce clinical situations of greater stress, demanding from the professional greater performance in the management of a child’s behavior. Such cases might require more restrictive techniques^7^. Therefore, dentists should pay particular attention to parents’ acceptance of BGT use to accomplish children’s treatment. It is noteworthy, however, that no scientific evidence is available to attest to the parents’ agreement with BGT. Thus, the purpose of this systematic review was to evaluate parental agreement with BGT during dental visits.

## MATERIAL AND METHODS

### Study design

The protocol of this systematic review was planned following the recommendations of the Preferred Reporting Items for Systematic Reviews and Meta-Analysis Protocols (PRISMA-P)^8^. It was registered in the International Prospective Register of Systematic Reviews (PROSPERO) under number CRD42018103834. The research is reported following the PRISMA Statement^9^.

### Study question

We addressed the acronym PECOS (Population, Exposition, Comparison, Outcomes, and Study design) to formulate the focused question: ‘What is the proportion of acceptance reported by the parents toward pediatric BGT?’ Where P – the parents of special health care needs (SHCN) children and the parents of non-special health care needs (non-SHCN) children submitted to dental care; E – the use of BGT in dental pediatric visits; C – none; O – the proportion of the parent’s acceptance with behavior guidance techniques; and S – observational studies.

### Eligibility criteria

To be included in this systematic review, the studies had to have observational designs. Studies that evaluated the parents’ agreement with BGT during the child’s dental treatment were included. Parents and legal guardians were accepted. The parents of non-special health care needs (non-SHCN) and special health care needs (SHCN) children of all ages were evaluated. Any kind of parental awareness of BGT (ex.: questionnaire, video, verbal or written information) was accepted. Due to limitation in publication records in some newer behavior guidance, the most BGT described by the AAPD in the current guideline^5^ were evaluated, including GA. Although hand over mouth (HOM) is no longer recommended by the guidelines, it was included in the study as well because many older studies have evaluated this technique. Hypnosis is not listed as one of the behavior management, also not in the past. It is worth mentioning, however, that primary studies did evaluate parents acceptance of hypnosis, therefore it was also evaluated. All dental procedures described in the studies were considered and all measures of the parents’ agreement were accepted.

The exclusion criteria were as follows: 1) Studies that did not evaluate the parents’ agreement of behavior guidance techniques but instead addressed the parents’ satisfaction/preferences and/or success rate and treatment costs, 2) Lacked data regarding parents’ agreement with BGT, 3) Secondary studies (review articles, letters to the editors, books, book chapters, etc.), 4) the studies that could not be found available in the complete text and 5) articles that duplicated participants from other publications.

### Information sources and search strategies

Detailed search strategies for each database were developed with the help of a health science librarian and they included MeSH terms and important synonyms (Appendix 1). The databases utilized were Cochrane Library, Latin American and Caribbean Health Sciences (LILACS), PubMed (including MedLine), PsycINFO, Scopus and Web of Science. A partial grey literature search was also carried out using the System for Information on the Grey Literature in Europe (OpenGrey), ProQuest Dissertations and Theses Database and Google Scholar. The search date was January 13^th^, 2019 and a search update was conducted on October 5^th^ 2020. No publication periods and language restrictions were applied. The reference lists from the included studies were also examined for relevant studies.

EndNote® X7 (Thomson Reuters, New York, EUA) and Rayyan software^10^ programs were used to manage the references. Duplicate identified studies were removed.

### Study selection and data collection process

Two reviewers (CM, JPS) independently selected the studies in two phases. First based on the titles and abstracts and in phase-two, based on the full-texts. The third reviewer (MB) made the final decision. The same procedure was applied for the meta-analysis data collection.

The following structured information was collected from each included study in pre-piloted forms: the authors, the year of publication, country, study design and setting, sample size, the participants’ gender, the children’s age, BGT, the measures of assessment of the BGT, main findings and the conclusions.

### Risk of bias in individual studies

The Joanna Briggs Institute Critical Appraisal Checklist for Analytical CrossLSectional Studies^11^ was used to assess the methodological quality of the individual included studies. The critical appraisal tool is composed of eight questions addressing the sample characteristics, the measurement of exposure, the condition being studied and any confounding factors. The possible answers to the tool’s questions are “yes”, if the study addressed the issue proposed in the question, “no” if the study did not address the issue, “unclear” in the case of unclear or information not completely reported; and “NA” for not applicable if a specific questions do not suit the issue addressed in the systematic review. The tool assesses the methodological quality of a study to determine extend to which it has addressed the possibility of bias in its design, conduct and analysis. The same two reviewers independently evaluated the included studies and disagreements were solved by consensus. As recommended by the reviewer’s manual, decisions about rating were discussed and agreed upon all reviewers before the critical appraisal begins. The grading system was determined by the authors considering: the studies that presented “yes” for all questions were rated as having good methodological quality therefore low risk of bias, those that presented at least one answer “unclear” was rated as unclear risk of bias, and at least one answer “no” was rated as high risk of bias. The plot was generated with the web app robvis^12^.

### Summary measures and synthesis of the results

The primary outcome was the proportion of the parent’s acceptance of BGT use for pediatric dental visits. Secondary outcomes included the differences in agreement with BGT between the parents of non-SHCN children and the parents of SHCN children and the differences in agreement with BGT between the parents who received an explanation before the presentation of the technique and those who did not. The proportion of the parent’s acceptance with the use of BGT was measured by a dichotomous outcome using the parent’s acceptance with each technique (yes/no) and the continuous outcome using the mean ratings of the parents’ agreement and the differences in means using a Visual Analog Scale (VAS) measured in millimeters (mm).

For data analysis, when the studies presented the mean VAS scores of the parents’ agreement using the rating anchors of zero mm as most accepted and 100 mm as the least accepted behavior technique, the data was transformed by reversing the value from 100 to zero to represent the least accepted and 100 mm to the most accepted. When the studies used a VAS measured in centimeters, the ratings were converted to mm. When the studies used a Likert scale, the “most acceptable” grade was pooled with the acceptance responses of “yes” for those studies that used “yes” or “no” for acceptance.

In addition, “conscious sedation” and “sedation” were pooled together as sedation, “parents’ separation” was combined with “parents present/ absent” and presented as “parental presence/absence”, “protective stabilization” and “physical restraints” were coded as active protective stabilization (APS) and “papoose board” and “passive restraint” were coded as passive protective stabilization (PPS).

Regarding SHCN children, independently of their specific health care needs, the parents’ agreement with BGT for all SHCN children were pooled together.

Studies with sufficient information were included in four different meta-analysis: 1) Proportion of acceptance with BGT separately for the parents of non-SHCN and SHCN children with the aid of MedCalc Statistical Software version 14.8.1 (MedCalc Software, Ostend, Belgium), 2) the mean of the agreement with BGT was measured with VAS for the parents of both non-SHCN children and SHCN children separately, with the aid of the Comprehensive Meta-Analysis Software (Biostat, Englewood, USA). All studies with the parents’ acceptance measured with VAS were included and a separate meta-analysis was performed for each BGT, 3) differences in the means of agreement with BGT measured with VAS among the parents of non-SHCN children were compared with the parents of SHCN children, using the RevMan Software (Review Manager, version 5.3, Cochrane Collaboration, Copenhagen, Denmark), and 4) differences in the means of agreement with BGT measured with VAS among the parents of non-SHCN who received an explanation before the presentation of the technique and those who did not, also measured with RevMan. Since the included studies were selected based on the inclusion and exclusion criteria, there was a potential for effects to be dissimilar, so a random-effects model was applied^13^. Heterogeneity was assessed using the I^2^ test (ratio of true heterogeneity to total observed variation) and a value >50% was considered to be an indicator of substantial heterogeneity between the studies^13^. The level of significance was set at 5%.

### Certainty of the evidence

Two independent reviewers (CM, JPS) assessed the certainty of evidence using the Grading of Recommendations, Assessment, Development and Evaluation (GRADE)^14^ criteria. Disagreements were resolved by consensus. The overall certainty of evidence is presented with a Summary of Findings (SoF) table, from GRADEpro software (McMaster University, Hamilton, Canada) (Appendix 7). Aspects such as risk of bias, inconsistency, indirectness, imprecision, and publication bias are causes to lower the certainty of the evidence and the presence of a large effect, dose response gradient and controlling of plausible confounders are causes of increasing it in observational studies. Certainty of evidence starts with low in observational studies and can be either upgraded or downgraded.

## RESULTS

### Study selection

The literature search identified 1633 citations across the six databases. After deduplication, 876 articles remained. An additional 14 studies were identified in the grey literature search, and after the article reference list examination and updated search. The full text of the 81 studies was accessed and 49 were found to meet the inclusion criteria for the review. From these, 41 contained sufficient information to allow for quantitative analysis. The detailed search and selection criteria are presented in Figure 1. The excluded studies with the exclusion rationale have been included in Appendix 2.

**Figure 1.**
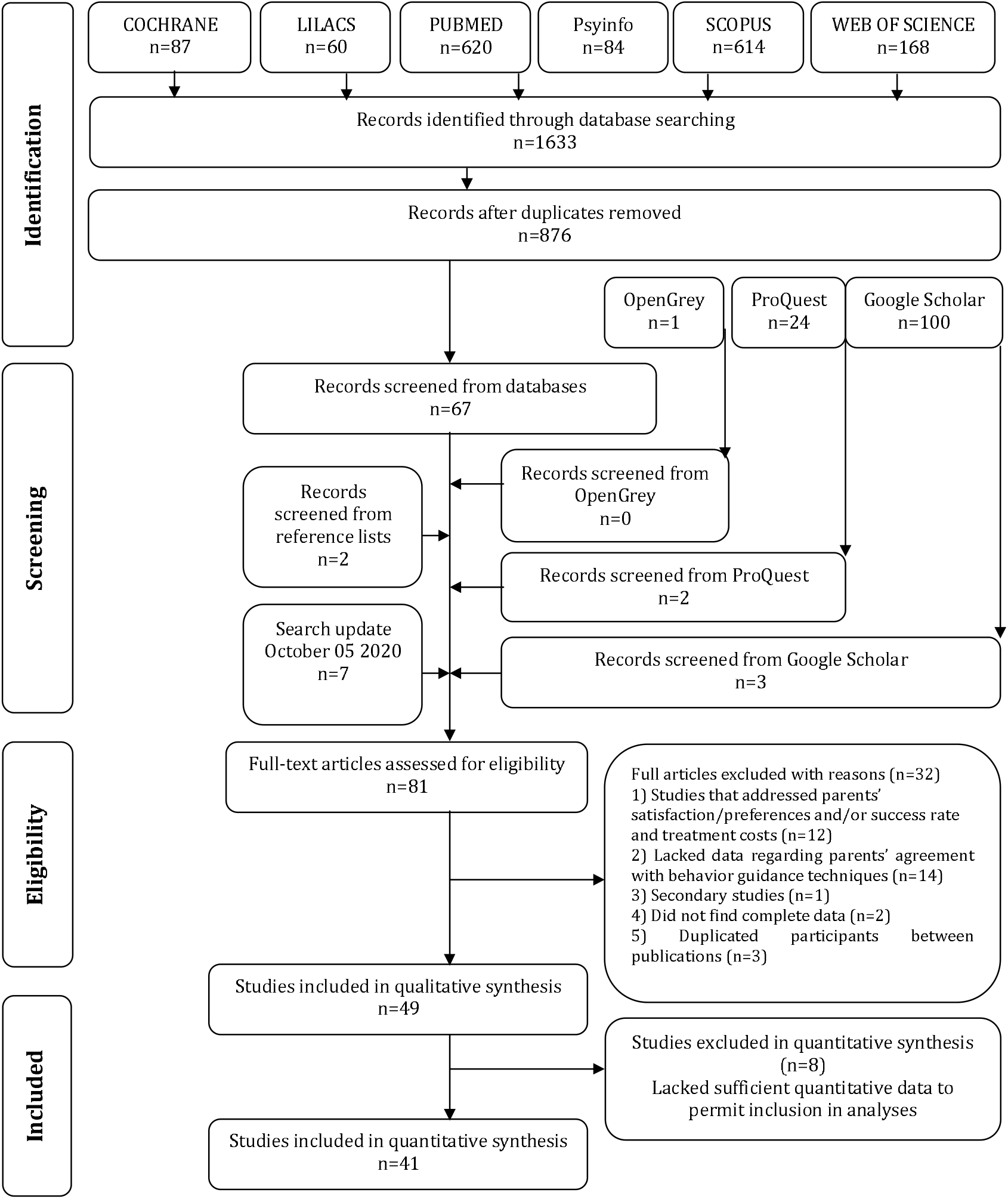
Flow diagram of literature search and selection criteria.^1^.

### Study characteristics

All of the 49 studies had cross-sectional designs with the enrollment of 4474 participants; and were published between 1984 and 2019. Most of the studies were conducted in clinics and pediatric hospitals (Table 1).

**Table 1.**
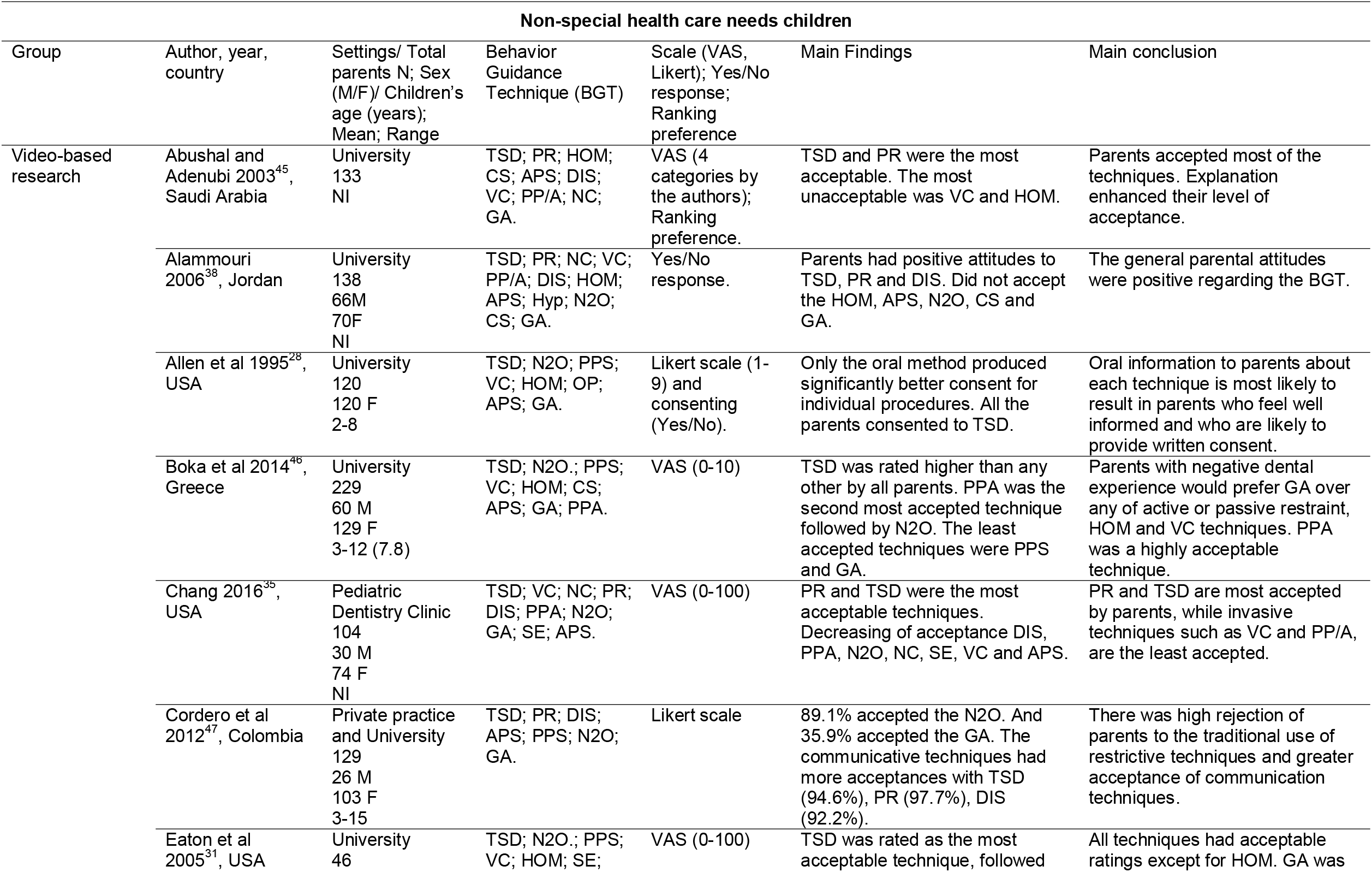
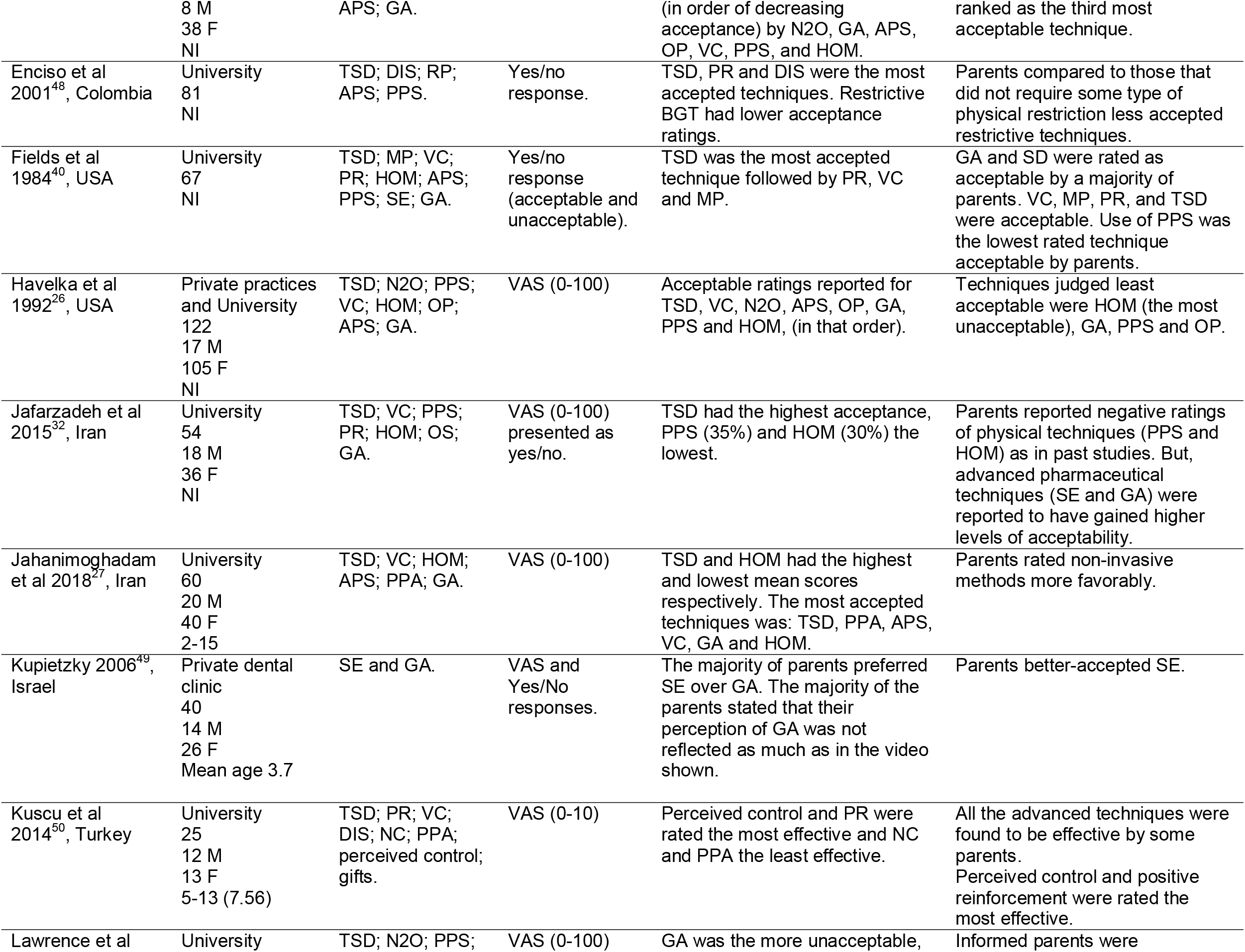
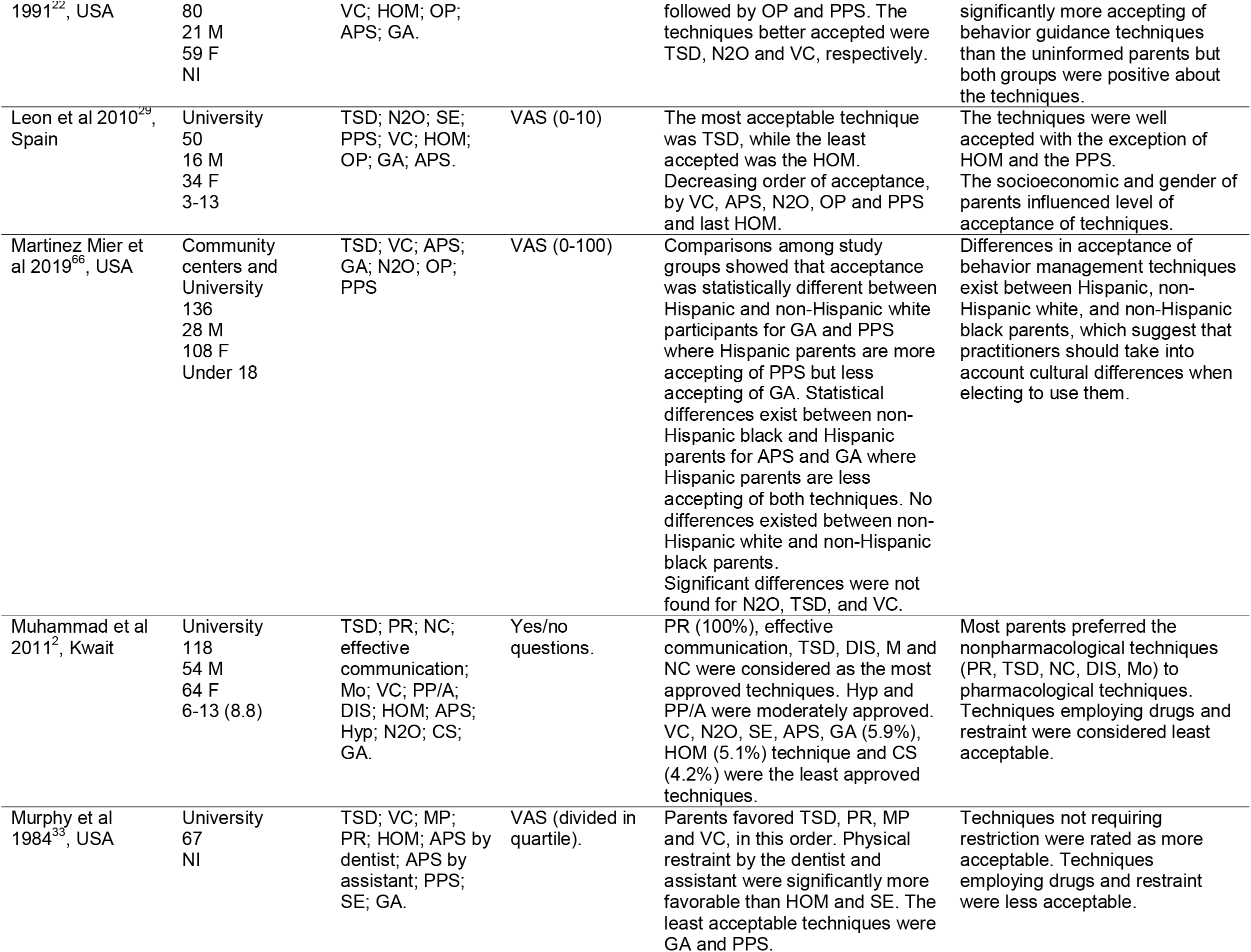
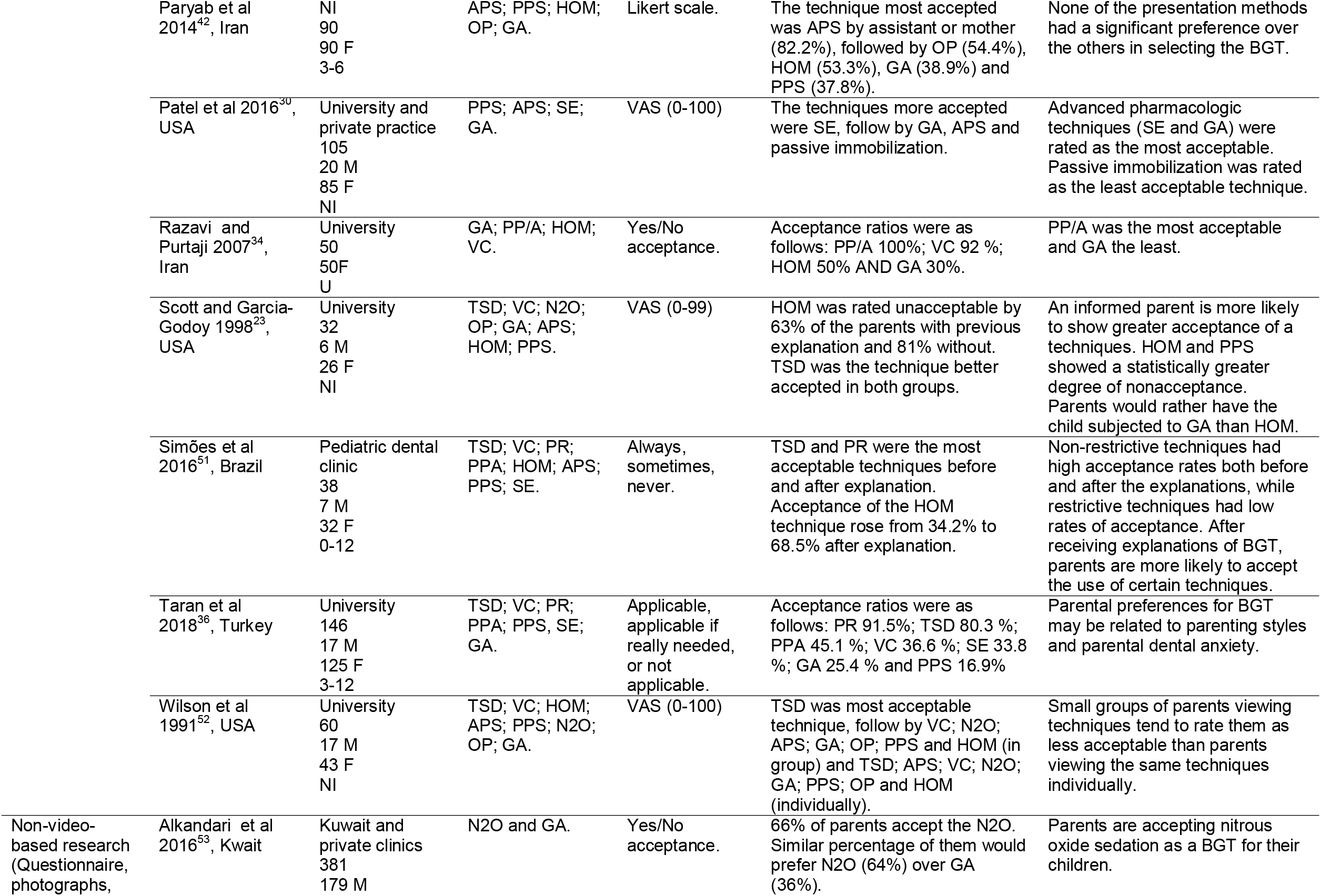
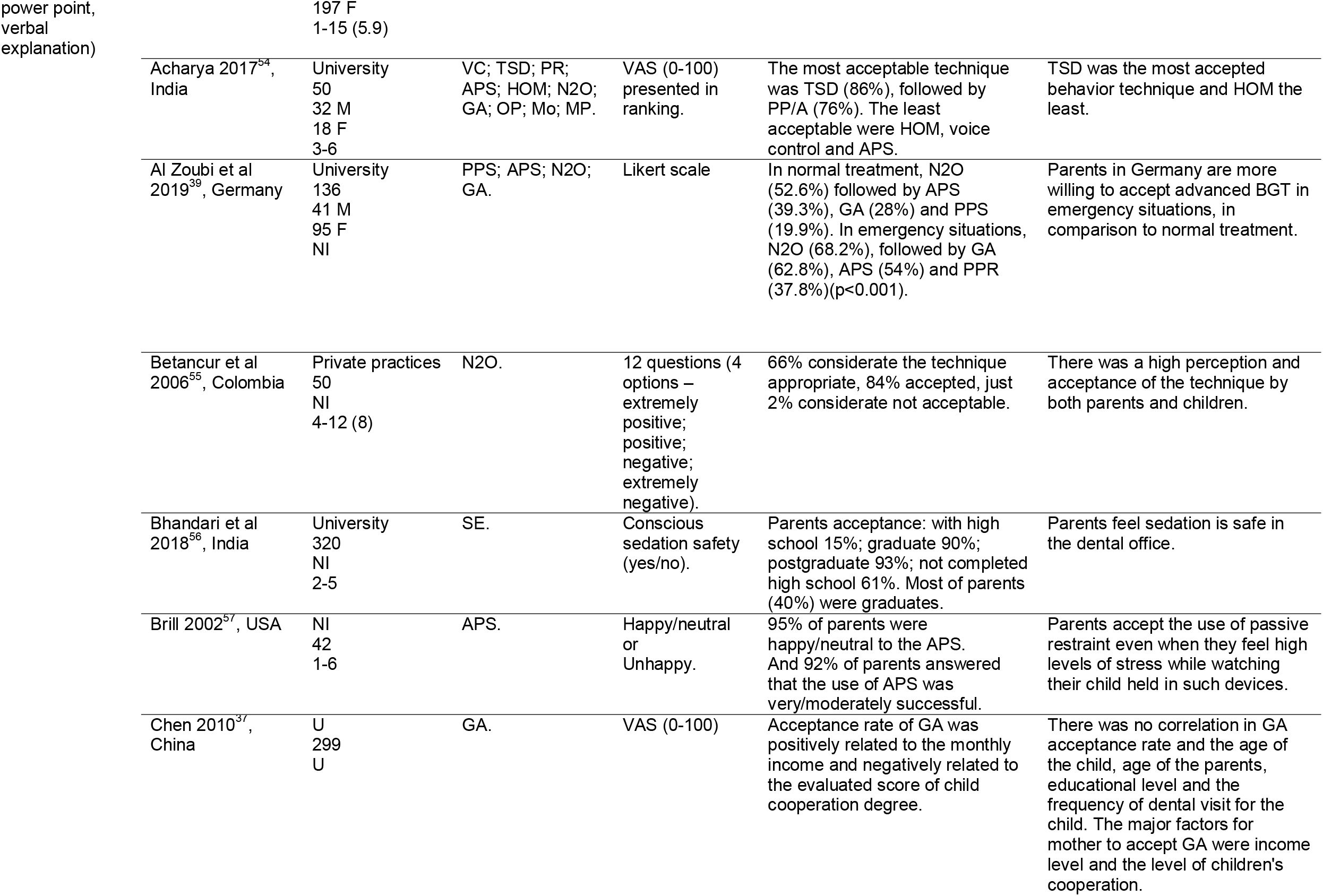
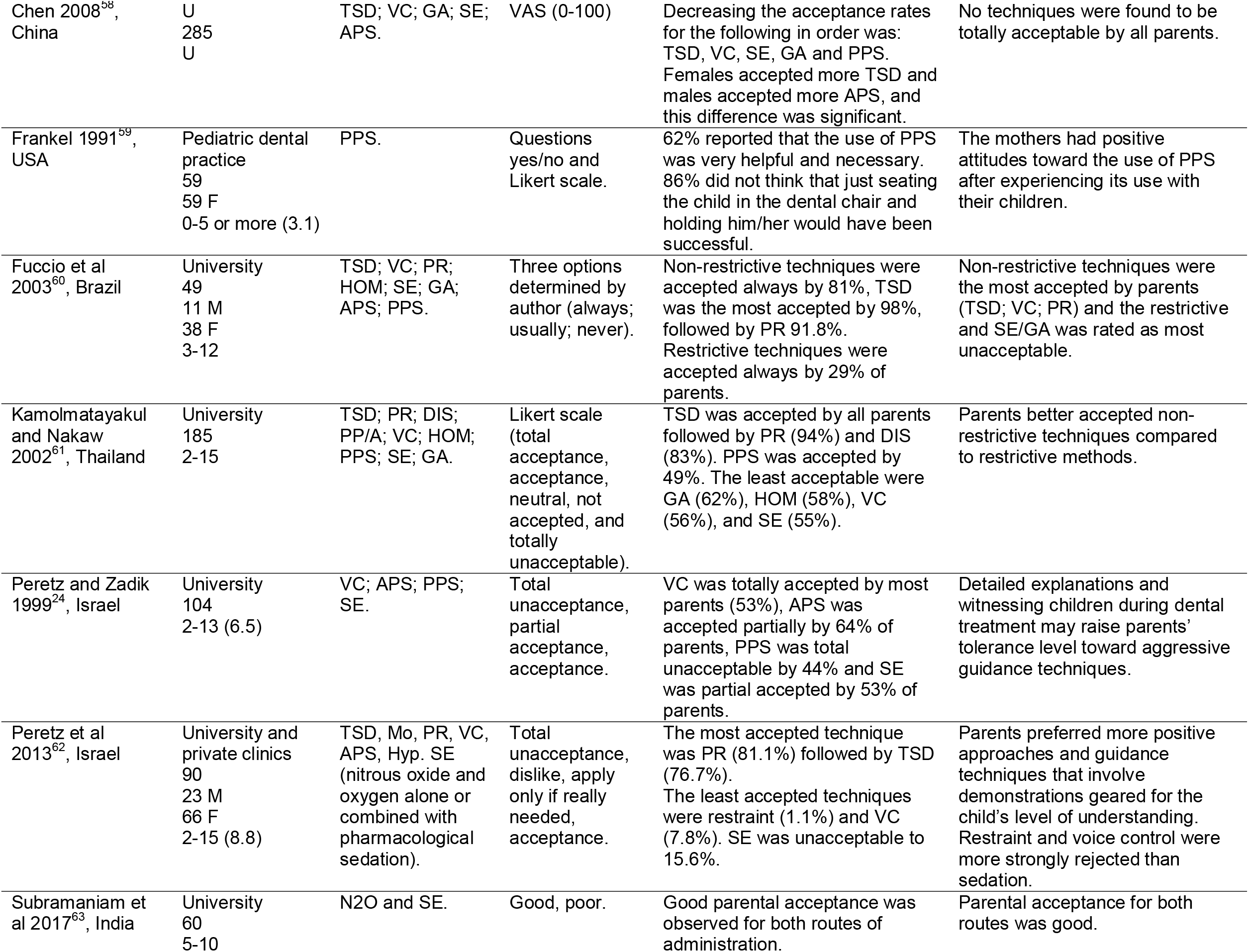
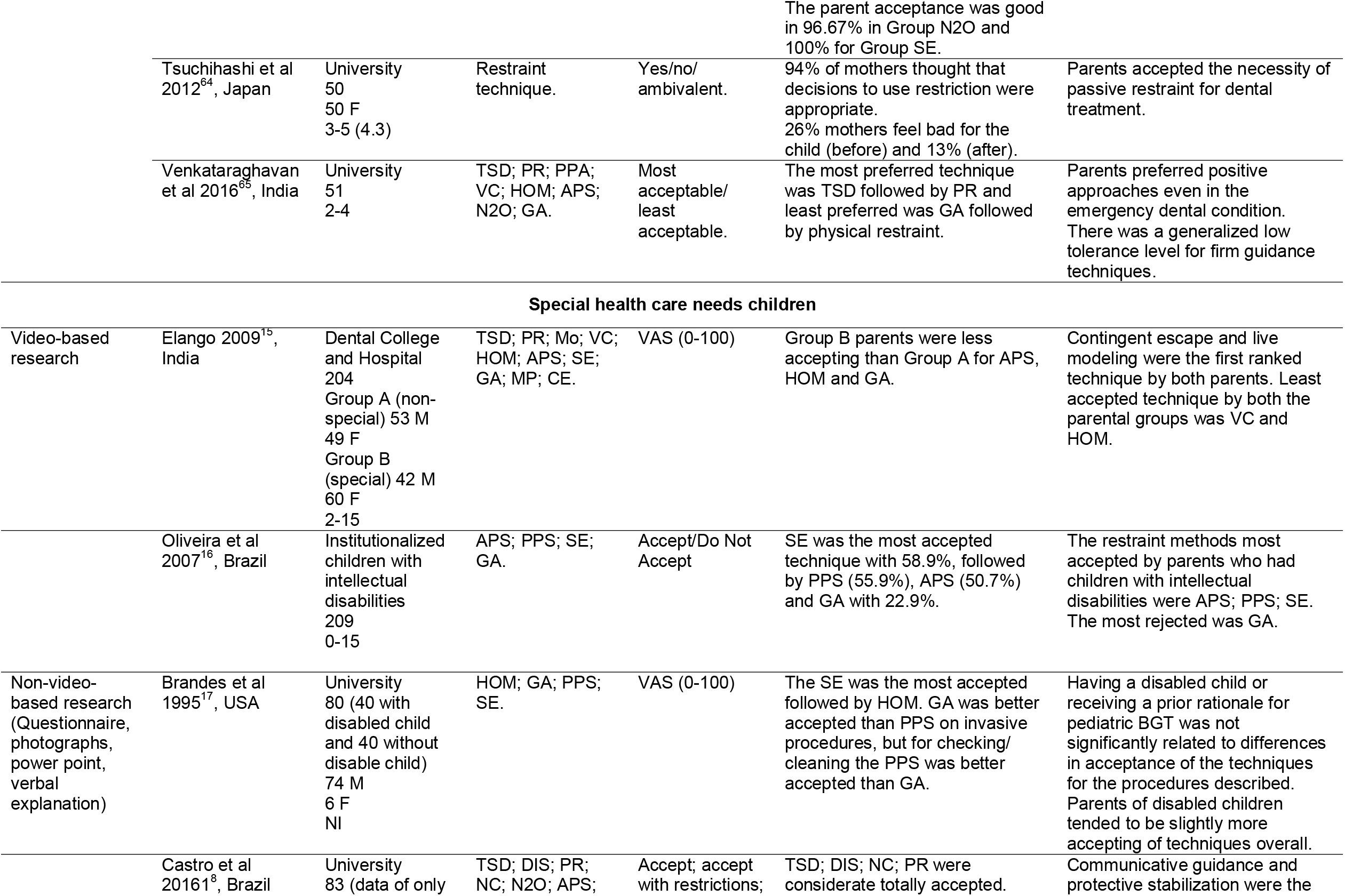
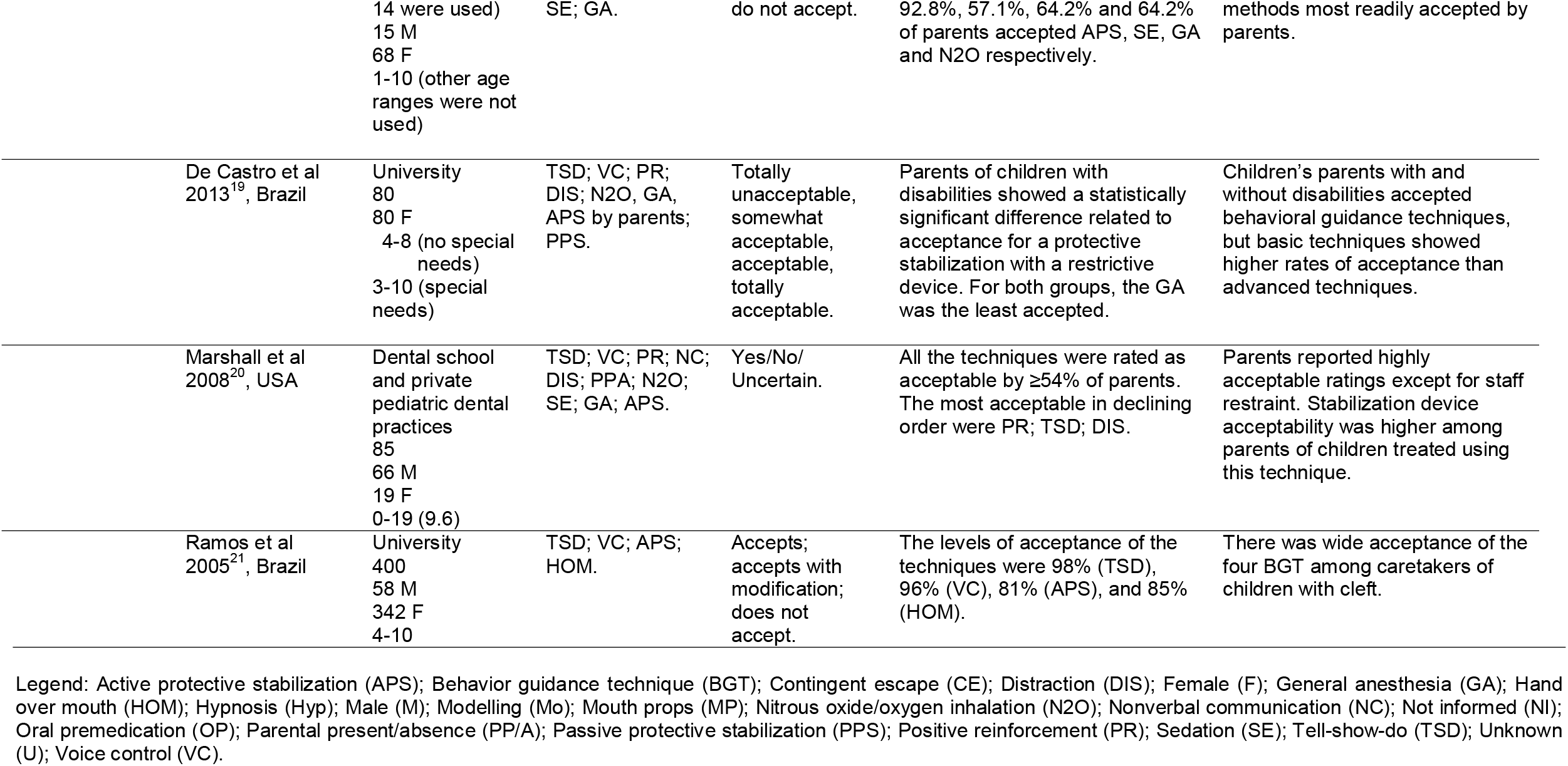
Summary of descriptive characteristics of included articles in non-special health care needs children and special health care needs children.

Seven studies evaluated the parents of SHCN children. The children were medically or physically compromised with neuropathological disorders^15^, with intellectual disabilities^16^, with physical or mental disabilities^17^, with physical or congenital disabilities, with mental, intelligence or behavioral deviations and/or systemic chronic diseases^18^ with a range of disabilities including Down’s Syndrome and cerebral palsy^19^. They also may have had autism^20^, and a cleft lip and/or palate ^21^ (Table 1).

### Risk of bias within the studies

The assessment of risk of bias is presented in Figure 2. According to Joanna Briggs Critical Appraisal Tool assessment, overall 33 studies were assessed as high risk of bias; five as unclear and only as 11 low risk of bias. A major concern regarding the risk of bias was observed, mainly issues with response rate, representativeness, and confounding.

**Figure 2.**
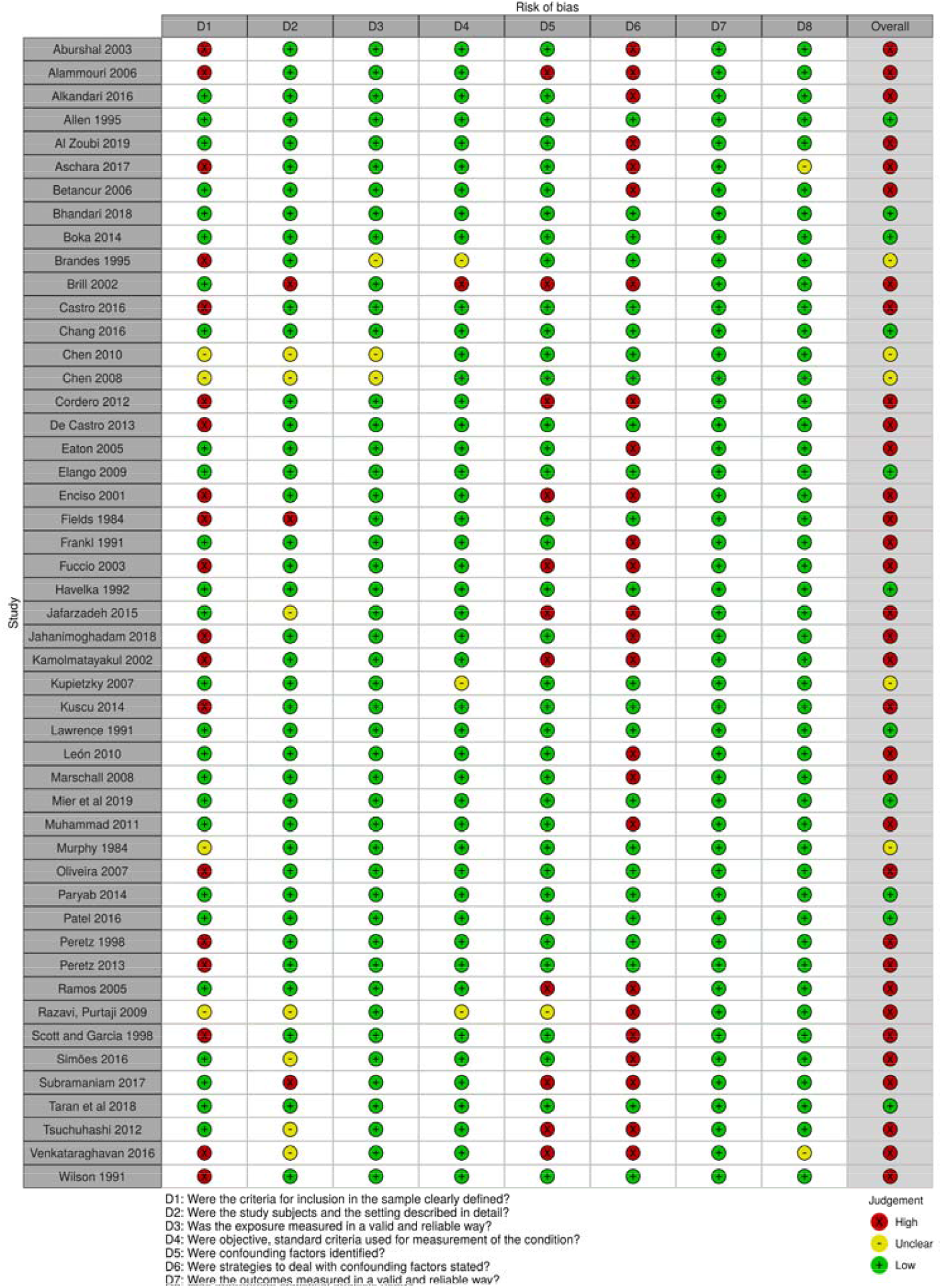
Methodological quality assessed by the Joanna Briggs Institute Critical Appraisal tools - Checklist for Analytical Cross-Sectional Studies. The studies that presented “yes” for all questions were rated as having low risk of bias, those that presented at least one answer “unclear” was rated as unclear risk of bias, and at least one answer “no” was rated as high risk of bias. Plot generated with the web app robvis.

### Synthesis of the results

The pooled analysis results for the primary outcome, namely the proportion of the parent’s agreement with the use of BGT for pediatric dental visits, were as follows:

1. The proportion of agreement with BGT by the parents of non-SHCN, reported based on acceptability/unacceptability, was examined using a separate meta-analysis for each technique. Overall, the analysis included 29 studies (n=2594) that evaluated 16 different BGT techniques. The random-effects model was employed. The proportion of acceptance varied from 84.1% (95%; confidence interval (CI) 75.8 to 90.9) to 21.2% (95% CI 11.0 to 33.7; p<0.001; I^2^ 94.5%) with TSD being found to be the most acceptable and HOM the least accepted (Figure 3 and Table 2). The I^2^ statistics, which refer to the proportion of the observed variance that reflects the differences in the true effects sizes (in log units)^13^, varied from not important at 32.5% (oral premedication) to considerable at 98.1% (modeling). Since I^2^ >50% was considered to be an indication of high heterogeneity, most of the meta-analysis showed considerable heterogeneity. The proportion of agreement with BGT by the parents of SHCN children analysis included five studies (n=748) with nine BGT techniques analyzed. The most accepted BGT in this analysis was TSD with 89.1% (95% CI 56.1 to 99.7; p<0.001; I^2^ 95.7%) of the parents agreeing and the least accepted was GA with 29.1% (95% CI 11.8 to 50.0; p=0.001; I^2^ 84.8). HOM was not assessed (Figure 4 and Table 3). The I^2^ statistics varied from zero (SE) to 98.5% (VC).
2. The mean of agreement with BGT measured with VAS for parents of non-SHCN children has been presented (Appendix 3). The random-effects model was employed. Distraction was the most accepted BGT with a mean of 94.2 mm (95% CI 93.6 to 94.8; p=0.423; I^2^ 0%) and PPS was the least accepted with the parents showing a mean of 42.2 mm (95% CI 29.4 to 55.0; p<0.001; I^2^ 99.8%) in VAS. The I^2^ varied from zero (TSD, PR, distraction, N_2_O, SE and GA) to 67.6% (PP/A).

**Table 2.** Proportion meta-analysis of agreement with BGT by the parents of non-SHCN children

| **Behavior Guidance**<br>**Technique** | **Total of**<br>**studies** | **Total of**<br>**sample** | **Proportion** | **CI 95%** | **p-value** | **I<sup>2</sup>** |
| --- | --- | --- | --- | --- | --- | --- |
| Tell-show-do | 15 | 1346 | 84.1% | 75.8-90.9 | <0.001 | 93.3 |
| Positive Reinforcement | 13 | 1188 | 82.1% | 73.3-89.3 | <0.001 | 92.4 |
| Distraction | 7 | 748 | 73.1% | 49.9-90.9 | <0.001 | 97.7 |
| Modeling | 6 | 474 | 67.6% | 34.8-92.8 | <0.001 | 98.1 |
| Nitrous oxide/oxygen<br>inhalation | 9 | 1062 | 59.1% | 38.5-78.2 | <0.001 | 97.6 |
| Mouth prop | 2 | 117 | 54.9% | 30.8-77.8 | 0.006 | 86.4 |
| Oral premedication | 4 | 194 | 50.1% | 41.5-58.6 | 0.227 | 32.5 |
| Nonverbal communication | 3 | 313 | 49.4% | 15.3-83.9 | <0.001 | 95.7 |
| Parental presence/absence | 7 | 732 | 49.2% | 26.3-72.3 | <0.001 | 97.6 |
| Voice control | 13 | 1082 | 40.2% | 24.4-57.2 | <0.001 | 96.8 |
| Active protective stabilization | 17 | 1333 | 38.7% | 25.0-53.3 | <0.001 | 96.6 |
| Sedation | 11 | 1313 | 33.7% | 18.1-51.9 | <0.001 | 97.7 |
| Hypnosis | 3 | 346 | 32.5% | 7.12-65.5 | <0.001 | 97.5 |
| General Anesthesia | 15 | 1681 | 27.4% | 16.8-39.4 | <0.001 | 96.3 |
| Passive protective stabilization | 12 | 1129 | 25.7% | 17.8-34.4 | <0.001 | 90.4 |
| Hand over mouth | 11 | 896 | 21.2% | 11.0-33.7 | <0.001 | 94.5 |

**Table 3.** Proportion meta-analysis of agreement with BGT by the parents of SHCN children

| **Behavior Guidance**<br>**Technique** | **Total of**<br>**studies** | **Total of**<br>**sample** | **Proportion** | **CI 95%** | **p-value** | **I<sup>2</sup>** |
| --- | --- | --- | --- | --- | --- | --- |
| Tell-show-do | 3 | 454 | 89.1% | 56.1-99.7 | <0.001 | 95.7 |
| Distraction | 2 | 54 | 83.4% | 32.5-98.4 | <0.001 | 92.6 |
| Positive reinforcement | 2 | 54 | 81.6% | 25.9-97.7 | <0.001 | 93.8 |
| Voice control | 2 | 440 | 73.8% | 12.5-98.1 | <0.001 | 98.5 |
| Active protective stabilization | 5 | 748 | 63.8% | 43.9-81.5 | <0.001 | 95.7 |
| Sedation | 2 | 223 | 58.6% | 52.1-65.0 | 0.871 | 0 |
| Passive protective stabilization | 3 | 334 | 47.2% | 33.0-61.6 | 0.003 | 82.6 |
| Nitrous oxide/oxygen<br>inhalation | 2 | 54 | 40.0% | 5.9-81.2 | 0.003 | 88.6 |
| General Anesthesia | 3 | 263 | 29.0% | 11.8-50.0 | 0.001 | 84.8 |

**Figure 3.**
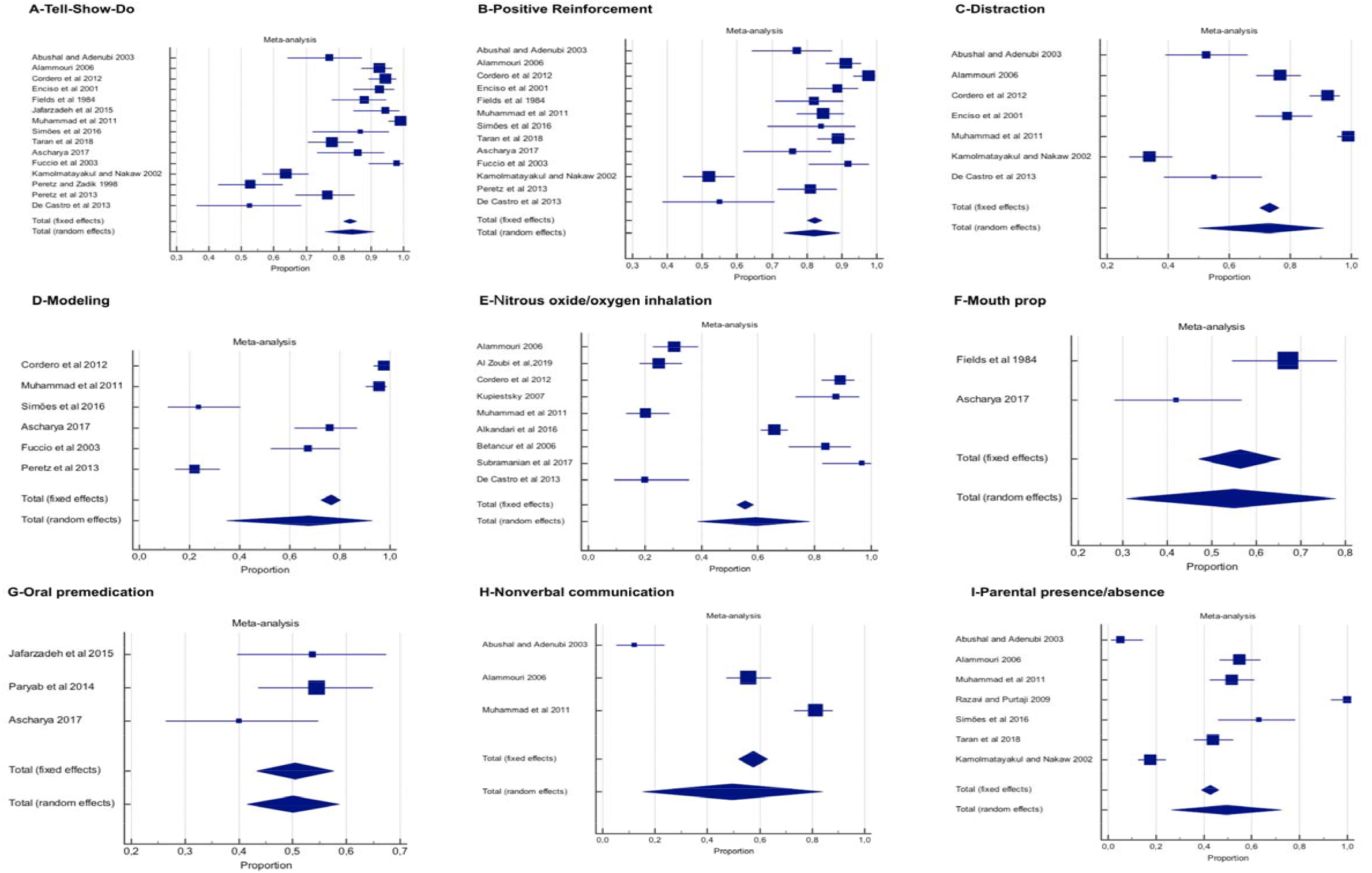
Meta-analysis of proportion (non-special heath care needs children)

**Figure 4.**
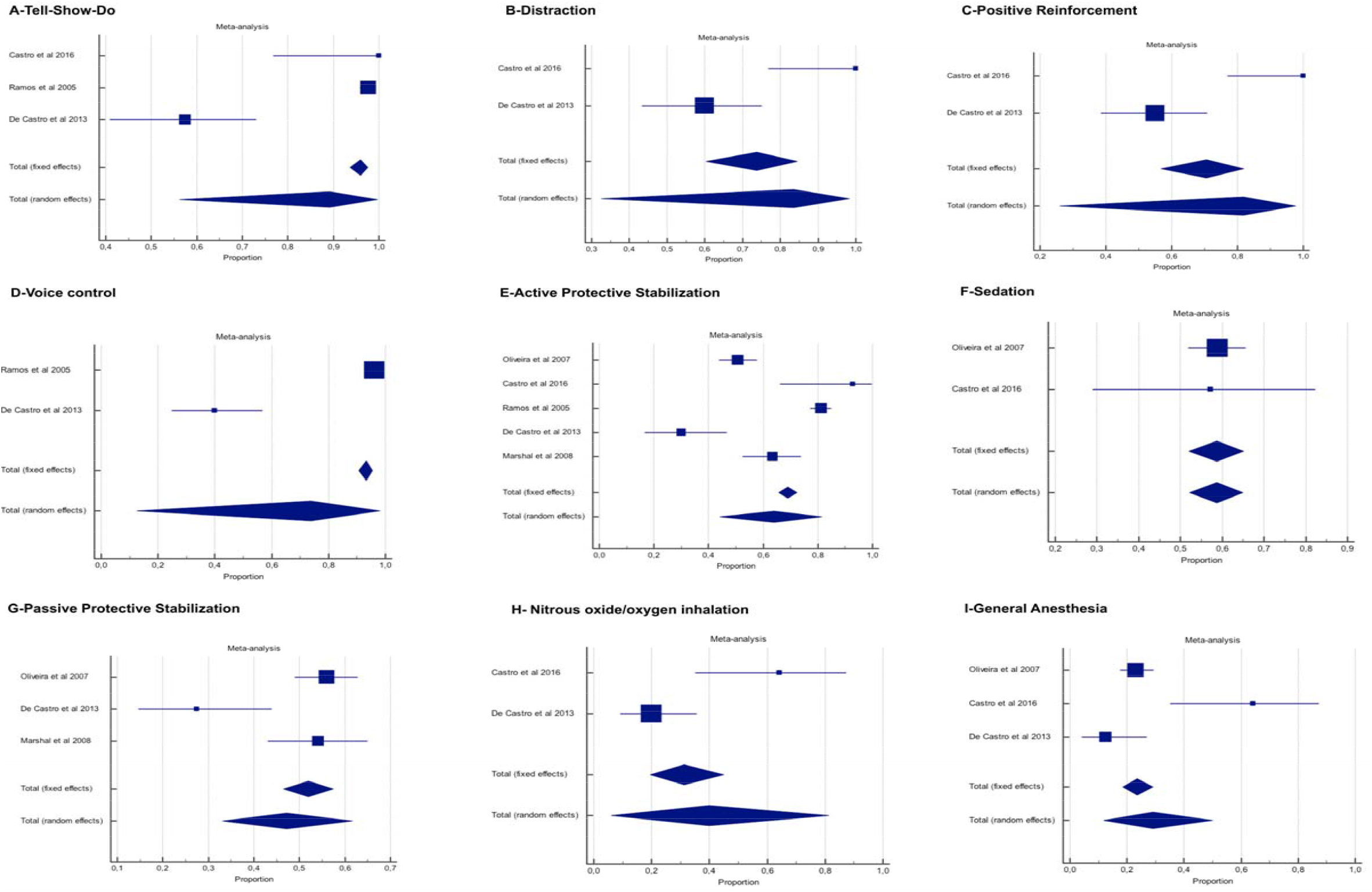
Meta-analysis of proportion special heath care needs children.

It was not possible to analyze the mean of the agreement with BGT measured with VAS for the parents of SHCN children due to the differences in the way that the data was presented among the studies.

The following meta-analyses show the results of the secondary outcomes:

1. The direct comparison of the acceptance of BGT among the parents of non-SHCN and SHCN children: the analyses were performed using two studies^15,17^ (n=245). The main outcome was the mean parental VAS rated acceptance in mm and the effect size was the standardized difference in mean. The random-effects model was again employed. The results showed that for active protective stabilization, the parents of SHCN children rated an average of 0.47 mm more for acceptance than the parents of non-SHCN children (Standard mean difference (SMD) 0.47; 95% CI 0.21 to 0.72; p<0.001; I^2^=0%). There was no significant difference found in the acceptance of HOM (SMD 0.22; 95% CI -0.03 to 0.47; p=0.08; I^2^=0%), SE (SMD 0.21; 95 % CI -0.04 to 0.46; p=0.10; I^2^=0%) and GA (SMD 0.07; 95% CI -0.18 to 0.32; p=0.57; I^2^=0%) (Appendix 4).
2. The difference in the means of an agreement with the BGT measured with VAS among the parents of non-SHCN children who received an explanation before the presentation of the technique and those who did not were examined. In the meta-analysis, the ratings from 112 parents from the two studies^22,23^ were made available. There was a significant difference in mean mm marked in VAS for those who received an explanation prior to judging the BGT for HOM (Mean difference (MD) -18.2; 95% CI -30.2 to -6.2; p=0.003; I^2^=94%); APS (MD -13.7; 95% CI -22.1 to -5.2; p=0.002; I^2^=89%) and TSD (MD - 9.8; 95% CI -12.7 to -7.0; p<0.001; I^2^=75%) with zero mm representing the most acceptable. The variable ‘had received an explanation’ did not significantly increase the parents’ agreement with N_2_O, GA, PPS, oral premedication and VC. A detailed analysis has been presented in Appendix 5. There was not enough data to analyze the parents of SHCN children.

### Results of the individual studies

A synthesis of parental acceptance and the scales used to measure it in the included studies are presented in Table 1. Overall, both parents of non-SHCN and SHCN children accepted communicative techniques and reported negative ratings on restrictive ones. Also, parents that were informed enhanced their level of acceptance of all techniques. Children’s age, parents’ previous experience in the dentist, sex, number of children, ethnicity, parenting style and income showed mixed results regarding parents preferences. While were parents’ age, education level, reason for children’s visit to the dentist, and children’s previous experience did not affect significantly parents’ level of acceptance.

### Certainty of the evidence

The certainty of the evidence according to the GRADE^15^ criteria was judged to be very low (Appendix 6). Major concerns were related to risk of bias (very serious) related to lack of definition of eligibility criteria and confounding factors; inconsistency (very serious) with heterogeneity above 50% and wide confidence intervals suggesting very low confidence in the estimated effect, and imprecision (serious) with less than 400 observation for continuous measures. Indirectness was not a concern. Publication bias was considered undetected because potential conflict of interest in the included studies was not observed. Furthermore, there was an effort to make a wide search including gray literature.

## DISCUSSION

Understanding parental acceptance toward BGT may have implication for planning children’s oral health treatment. In the present systematic review, we found that parents of non-SHCN and SHCN children demonstrated high acceptance of basic behavior guidance. Regarding advanced behavior guidance, the proportion of acceptance was good among parents of SHCN children and low among parents of non-SHCN. Active protective stabilization was more accepted among special parents than among non-SHCN. Overall, explanation about the technique increased parental acceptance, however not for all the techniques. Nevertheless, the high risk o bias of the included studies; the high clinical, methodological and statistical heterogeneity; and the very low certainty of the evidence represent a challenge in interpreting the results.

Perhaps the parents of SHCN children are more used to physical restraint, especially when their children present with aggressive behavior^16^. This could be the reason in the results as to why the parents accept protective stabilization and sedation leaving N_2_O and GA as the last choices. Additionally, the parents of uncooperative SHCN children were more open to accepting advanced BGT ^20,24^.

For dental care providers, there is an obligation to offer accurate information to parents about their children’s treatment. In the case of need for advanced behavior guidance, dentists should support the decisions on the evidence-based guidelines and systematic reviews. Nevertheless, the potential harm of a more invasive guidance technique such as protective stabilization or GA should be considered along with parents’ opinions^5^. A 2-way conversation about risks and benefits of BGT allows parents to express their values and preferences while sharing the choice with the oral care team regarding the best way their children could be treated^25^. Moreover, well-informed parents accept better^26,27^ and are more prone to give consent on BGT use^28^.

Children present multifaceted behavior according to their age range. The present study analysis did not approach parents’ BGT acceptance regarding children’s age because there was not sufficient homogeneous data to perform subgroup analysis among included studies. However, studies showed mixed results suggesting that age did not affect significantly parents’ level of acceptance^7^. In other case younger ages presented greater parents’ acceptability to N_2_O^28^. Likewise, parents’ previous experience in the dentist^29,30^, sex^2,31,32,29^, number of children^33,34^, ethnicity^2,35^, parenting style^36,24^ and income^37,31,32,2,29,33^ showed controversial results while parents’ age^31,32^, education level^2,31,32^, reason for children’s visit to the dentist^7^, and children’s previous experience^38,29^ did not affect significantly parents’ level of acceptance. Unfortunately, there is no reliable anticipatory way dentists can predict which BGT will be more likely to be accepted.

The results, however, allowed to observe that in cases of pain and/or emergency and uncooperative children, parents were more willing to accept advanced techniques^24,39,40,30^. Furthermore, parents of cooperative children did not approve sedation^24^ while stressed parents accepted less BGT^22^. Therefore, recommendations would rely on using the technique that can provide the behavior management that is particularly needed to effectively treat the child. Usually, dentists pay attention to the parent-child relationship; therefore the results of the present review may help dentists to seek for the parent acceptance of the more suitable BGT for that particular family.

Different relationships may be obtained in different countries. Culture and social mores can influence on the parents point of view in the dental visit approach. Each country has state laws and regulations concerning dental practice and BGT are included in these regulatory efforts. For instance, in Nordic European countries, devices for protective stabilization are forbidden^41^. Advanced behavior guidance requires informed consent signed by the parents and kept in the patient record^6^. Even when basic behavior techniques are planned, informed consent is required for alternative methods in case of the necessity to change the BGT^41^.

Although HOM is a technique no longer accepted, it was included in the present systematic review due to the number of the included studies that have assessed it. Indeed parents showed disagreement about the use of HOM. There are growing concerns regarding the ethical boundaries of more restrictive techniques^42,43^ especially if the dentist does not have the scientific knowledge and training to perform it^5^. Even for SHCN children that present limited cooperation, physical restraint is seen as a final option for managing behavior^44^.

This systematic review also investigated hypnosis. The agreement with hypnosis varied from low^24^ to moderate^2^. The parents that agreed were more likely to be women^38^, older and younger children^24^. Perhaps parents’ perceptions of the benefits to the child anxiety favor their acceptance of the technique.

There are common issues among the included studies that compromise the present results. Firstly, most of them did not present inclusion criteria, did not present sample size calculation, did not describe the settings and did not address confounding factor such as participants’ age, socioeconomic characteristics, previous experience with the dentist and with BGT, number of siblings, anxiety, pain and treatment. Secondly, the methodological problems certainly affect solid conclusions. Another limitation is the outcome measurement. The included studies used a range of scales to access parents acceptance with a range of methods to present BGT to parents.

The SHCN children were assessed without any differences in their health conditions and the limitations associated with those conditions. It is possible that the parents’ acceptance would be different among the children with a condition such as cerebral palsy, especially because the parents are used to stabilization depending on the level of the disability when compared with the parental preferences for children with systemic chronic diseases. Furthermore, some health disabilities were not assessed such as deafness and blindness.

The present systematic review had a comprehensive search including grey literature with the help of a health science librarian, and presented a high number of include studies, however it is not possible to be sure that all possible eligible studies were included. Also, the effect estimates varied greatly since substantial heterogeneity across studies was observed limiting the confidence in the results. All the mentioned limitation influenced the GRADE assessment, which showed very low-level certainty of the overall evidence.

Based on the issues herein discussed, it is clear that all the pointed limitations affect the present systematic review conclusions and applicability. Yet, dentists should discuss BGT options with parents having in mind that generally basic guidance techniques are well accepted among parents of non-SHCN children as well as among parents of SHCN and probably, for advanced behavior guidance, there will be more resistance among all parents. Moreover, explanation increased parents’ acceptability.

Future research should address the BGT presented in the current AAPD guideline^5^ such as positive pre-visit imagery, ask-tell-ask, memory recruiting and the communication techniques for parents, which involves ask-tell-ask, teach-back and motivational interviewing.

## Conclusions

This systematic review and meta-analysis suggests with very low certainty that parents’ attitudes towards BGT are more likely to be accepting of basic behavior guidance with a high level of acceptance and less likely to accept advanced behavior guidance. This was the case for both parents of non-SHCN and SHCN children. Parents were less likely to accept more restrictive measures. Further, there is some evidence that parents’ benefit from education and experience with respect to BGT suggesting that dentists should discuss BGT options with both the parents of non-SHCN and SHCN children. These findings provide potentially helpful direction for dental care providers aimed at improving child health and child- and family-centered dental care.

## Data Availability

All data are available by sending an email to

## Acknowledgments

Thanks to Mrs. Maria Gorete Savi for her contribution in the search strategy.

## Footnotes

1 Adapted from PRISMA.

