## Supplemental Tables and Figures for "Parental acceptance toward behavior guidance techniques for pediatric dental visits: a meta-analysis"

**Appendix 1** **-** Search Strategy.

| **Database** | **Search** |
| --- | --- |
| **Cochrane** | ((("Conditioning, Operant" OR "Operant Conditioning" OR "Operant Conditionings" OR "Instrumental Learning" OR "Restraint, Physical" OR "Physical Restraint" OR "Physical Restraints" OR "Physical Immobilization" OR "Immobilization" OR "Persuasive Communication" OR "Conscious Sedation" OR "Reinforcement (Psychology)" OR "Reinforcement" OR "Reinforcements" OR "Collaboration" OR "Collaborations" OR "co-operation" OR "co-operations" OR "cooperation" OR "cooperations" OR "Accepting" OR "acceptance" OR "Behavior Control" OR "Behavior Therapy" OR "Problem Behavior" OR "Cooperative Behavior") AND ("parents" OR "parent" OR "Parent-Child Relations" OR "parental" OR "mothers" OR "mother" OR "fathers" OR "father") AND ("child" OR "children" OR "childhood" OR "child, preschool" OR "preschool" OR "preschools" OR "pediatrics" OR "pediatric" OR "paediatrics" OR "paediatric" OR "Child Behavior") AND ((("dental" OR "dentistry") AND ("visit" OR "visits" OR "treatment" OR "treatments" OR "restoration" OR "restorations" OR "Tooth Extraction" OR "Extraction" OR "Extractions" OR "Dental Prophylaxis" OR "Prophylaxis")) OR "Dental Care" OR "Dental Care for Children" OR "Dental Offices" OR "Dental Office" OR "Pediatric Dentistry" OR "oral health") |
| **LILACS** | (tw:(((("dental" OR "dentistry") AND ("visit" OR "visits" OR "treatment" OR "treatments" OR "restoration" OR "restorations" OR "Tooth Extraction" OR "Extracción Dental" OR "Extração Dentária" OR "Extraction" OR "Extractions" OR "Dental Prophylaxis" OR "Prophylaxis")) OR "Dental Care" OR "Dental Care for Children" OR "Dental Offices" OR "Dental Office" OR "Pediatric Dentistry" OR "Odontología Pediátrica" OR "Odontopediatria" OR "oral health"))) AND (tw:(("Conditioning, Operant" OR "Operant Conditioning" OR "Operant Conditionings" OR "Condicionamiento Operante" OR "Condicionamento Operante" OR "Instrumental Learning" OR "Restraint, Physical" OR "Physical Restraint" OR "Physical Restraints" OR "Physical Immobilization" OR "Restricción Física" OR "Restrição Física" OR "Immobilization" OR "Persuasive Communication" OR "Comunicación Persuasiva" OR "Comunicação Persuasiva" OR "Conscious Sedation" OR "Sedación Consciente" OR "Sedação Consciente" OR "Reinforcement(Psychology)" OR "Refuerzo (Psicología)" OR "Reforço (Psicologia)" OR "Reinforcement" OR "Reinforcements" OR "Reinforcement, Verbal" OR "Refuerzo Verbal" OR "Reforço Verbal" OR "Collaboration" OR "Collaborations" OR "co-operation" OR "co-operations" OR "cooperation" OR "cooperations" OR "Accepting" OR "acceptance" OR "Patient Acceptance of Health Care" OR "Aceptación de la Atención de Salud" OR "Aceitação pelo Paciente de Cuidados de Saúde" OR "Behavior Control" OR "Behavior Therapy" OR "Problem Behavior" OR "Cooperative Behavior" OR "Conducta Cooperativa" OR "Comportamento Cooperativo"))) AND (tw:(("parents" OR "Padres" OR "Pais" OR "parent" OR "Parent-Child Relations" OR "Relaciones Padres-Hijo" OR "Relações Pais-Filho" OR "parental" OR "mothers" OR "madres" OR "mães" OR "mother" OR "madre" OR "mãe" OR "fathers" OR "father" OR "padre" OR "pai") )) AND (tw:(("child" OR "Niño" OR "criança" OR "children" OR "childhood" OR "child, preschool" OR "preschool" OR "Preescolar" OR "Pré-Escolar" OR "preschools" OR "pediatrics" OR "pediatric" OR "paediatrics" OR "paediatric" OR "Child Behavior"))) AND (instance:"regional") AND ( db:("LILACS")) |
| **PsycInfo** | (((Any Field: "Conditioning, Operant" OR Any Field: "Operant Conditioning" ORAny Field: "Operant Conditionings" OR Any Field: "Instrumental Learning" OR Any Field: "Restraint, Physical" OR Any Field: "Physical Restraint" OR Any Field: "Physical Restraints" ORAny Field: "Physical Immobilization" OR Any Field: "Immobilization" OR Any Field: "Persuasive Communication" OR Any Field: "Conscious Sedation" OR Any Field: "Reinforcement(Psychology)" OR Any Field: "Reinforcement" OR Any Field: "Reinforcements" ORAny Field: "Collaboration" OR Any Field: "Collaborations" OR Any Field: "co-operation" OR Any Field: "co-operations" OR Any Field: "cooperation" OR Any Field: "cooperations" OR Any Field: "Accepting" OR Any Field: "acceptance" OR Any Field: "Behavior Control" OR Any Field: "Behavior Therapy" OR Any Field: "Problem Behavior" OR Any Field: "Cooperative Behavior")AND (Any Field: "parents" OR Any Field: "parent" OR Any Field: "Parent-Child Relations" ORAny Field: "parental" OR Any Field: "mothers" OR Any Field: "mother" OR Any Field: "fathers"OR Any Field: "father") AND (Any Field: "child" OR Any Field: "children" OR Any Field: "childhood" OR Any Field: "child, preschool" OR Any Field: "preschool" OR Any Field: "preschools" OR Any Field: "pediatrics" OR Any Field: "pediatric" OR Any Field: "paediatrics" ORAny Field: "paediatric" OR Any Field: "Child Behavior") AND (((Any Field: "dental" OR Any Field: "dentistry") AND (Any Field: "visit" OR Any Field: "visits" OR Any Field: "treatment" OR Any Field: "treatments" OR Any Field: "restoration" OR Any Field: "restorations" OR Any Field: "Tooth Extraction" OR Any Field: "Extraction" OR Any Field: "Extractions" OR Any Field: "Dental Prophylaxis" OR Any Field: "Prophylaxis")) OR Any Field: "Dental Care" OR Any Field: "Dental Care for Children" OR Any Field: "Dental Offices" OR Any Field: "Dental Office" OR Any Field: "Pediatric Dentistry" OR Any Field: "oral health") AND Document Type: Journal Article |
| **PubMed** | ((("Conditioning, Operant"[Mesh] OR "Operant Conditioning"[All Fields] OR "Operant Conditionings"[All Fields] OR "Instrumental Learning"[All Fields] OR "Restraint, Physical"[Mesh] OR "Physical Restraint"[All Fields] OR "Physical Restraints"[All Fields] OR "Physical Immobilization"[All Fields] OR "Immobilization"[Mesh] OR "Immobilization"[All Fields] OR "Persuasive Communication"[Mesh] OR "Persuasive Communication"[All Fields] OR "Conscious Sedation"[Mesh] OR "Conscious Sedation"[All Fields] OR "Reinforcement(Psychology)"[Mesh:noexp] OR "Reinforcement"[All Fields] OR "Reinforcements"[All Fields] OR "Collaboration"[All Fields] OR "Collaborations"[All Fields] OR "co-operation"[All Fields] OR "co-operations"[All Fields] OR "cooperation"[All Fields] OR "cooperations"[All Fields] OR "Accepting"[All Fields] OR "acceptance"[All Fields] OR "Behavior Control"[Mesh] OR "Behavior Therapy"[Mesh] OR "Problem Behavior"[Mesh] OR "Cooperative Behavior"[Mesh] OR "Behavior Control"[All Fields] OR "Behavior Therapy"[All Fields] OR "Problem Behavior"[All Fields] OR "Cooperative Behavior"[All Fields] OR "cognitive therapy"[All Fields] OR "play therapy"[All Fields] OR "music therapy"[All Fields]) AND ("parents"[MeSH] OR "parents"[All Fields] OR "parent"[All Fields] OR "Parent-Child Relations"[Mesh] OR "parental"[All Fields] OR "mothers"[MeSH] OR "mothers"[All Fields] OR "mother"[All Fields] OR "fathers"[MeSH] OR "fathers"[All Fields] OR "father"[All Fields])) AND ("child"[MeSH Terms] OR "child"[Title/Abstract] OR "children"[Title/Abstract] OR "childhood"[Title/Abstract] OR "child, preschool"[MeSH Terms] OR preschool[All Fields] OR preschools[All Fields] OR "pediatrics"[MeSH Terms] OR "pediatrics"[Title/Abstract] OR "pediatric"[Title/Abstract] OR "paediatrics"[Title/Abstract] OR "paediatric"[Title/Abstract] OR "Child Behavior"[Mesh])) AND ((("dental"[Title/Abstract] OR "dentistry"[Title/Abstract]) AND ("visit"[All Fields] OR "visits"[All Fields] OR "treatment"[All Fields] OR "treatments"[All Fields] OR "restoration"[All Fields] OR "restorations"[All Fields] OR "Tooth Extraction"[Mesh:noexp] OR "Extraction"[All Fields] OR "Extractions"[All Fields] OR "Dental Prophylaxis"[Mesh:noexp] OR "Prophylaxis"[All Fields])) OR "Dental Care"[Mesh:noexp] OR "Dental Care"[All Fields] OR "Dental Care for Children"[Mesh] OR "Dental Offices"[Mesh] OR "Dental Offices"[All Fields] OR "Dental Office"[All Fields] OR "Pediatric Dentistry"[Mesh] OR "oral health"[Title/Abstract]) |
| **Scopus** | TITLE-ABS KEY ( ( ( "dental" OR "dentistry" ) AND ( "visit" OR "visits" OR "treatment" OR "treatments" OR "restoration" OR "restorations" OR "Tooth Extraction" OR "Extraction" OR "Extractions" OR "Dental Prophylaxis" OR "Prophylaxis" ) ) OR "Dental Care" OR "Dental Care for Children" OR "Dental Offices" OR "Dental Office" OR "Pediatric Dentistry" OR "oral health" ) AND TITLE-ABS-KEY ( ( "child" OR "children" OR "childhood" OR "child, preschool" OR "preschool" OR "preschools" OR "pediatrics" OR "pediatric" OR "paediatrics" OR "paediatric" OR "Child Behavior" ) ) AND TITLE-ABS-KEY ( ( "parents" OR "parent" OR "Parent-Child Relations" OR "parental" OR "mothers" OR "mother" OR "fathers" OR "father" ) ) AND TITLE-ABS-KEY ( ( "Conditioning, Operant" OR "Operant Conditioning" OR "Operant Conditionings" OR "Instrumental Learning" OR "Restraint, Physical" OR "Physical Restraint" OR "Physical Restraints" OR "Physical Immobilization" OR "Immobilization" OR "Persuasive Communication" OR "Conscious Sedation" OR "Reinforcement(Psychology)" OR "Reinforcement" OR "Reinforcements" OR "Collaboration" OR "Collaborations" OR "co-operation" OR "co-operations" OR "cooperation" OR "cooperations" OR "Accepting" OR "acceptance" OR "Behavior Control" OR "Behavior Therapy" OR "Problem Behavior" OR "Cooperative Behavior" ) ) AND ( LIMIT-TO ( DOCTYPE , "ar" ) ) |
| **Web of Science** | ((("Conditioning, Operant" OR "Operant Conditioning" OR "Operant Conditionings" OR "Instrumental Learning" OR "Restraint, Physical" OR "Physical Restraint" OR "Physical Restraints" OR "Physical Immobilization" OR "Immobilization" OR "Persuasive Communication" OR "Conscious Sedation" OR "Reinforcement(Psychology)" OR "Reinforcement" OR "Reinforcements" OR "Collaboration" OR "Collaborations" OR "co-operation" OR "co-operations" OR "cooperation" OR "cooperations" OR "Accepting" OR "acceptance" OR "Behavior Control" OR "Behavior Therapy" OR "Problem Behavior" OR "Cooperative Behavior") AND ("parents" OR "parent" OR "Parent-Child Relations" OR "parental" OR "mothers" OR "mother" OR "fathers" OR "father") AND ("child" OR "children" OR "childhood" OR "child, preschool" OR "preschool" OR "preschools" OR "pediatrics" OR "pediatric" OR "paediatrics" OR "paediatric" OR "Child Behavior") AND ((("dental" OR "dentistry") AND ("visit" OR "visits" OR "treatment" OR "treatments" OR "restoration" OR "restorations" OR "Tooth Extraction" OR "Extraction" OR "Extractions" OR "Dental Prophylaxis" OR "Prophylaxis")) OR "Dental Care" OR "Dental Care for Children" OR "Dental Offices" OR "Dental Office" OR "Pediatric Dentistry" OR "oral health") **Refined by:** **DOCUMENT TYPES:** ( ARTICLE ) |
| **Google Scholar** | (("parental" OR "mothers" OR "mother" OR "fathers" OR "father") AND ("acceptance")) AND (("child" OR "children") AND "dental" AND ("Behavior Control")) |
| **OpenGrey** | ((("Conditioning, Operant" OR "Operant Conditioning" OR "Operant Conditionings" OR "Instrumental Learning" OR "Restraint, Physical" OR "Physical Restraint" OR "Physical Restraints" OR "Physical Immobilization" OR "Immobilization" OR "Persuasive Communication" OR "Conscious Sedation" OR "Reinforcement(Psychology)" OR "Reinforcement" OR "Reinforcements" OR "Collaboration" OR "Collaborations" OR "co-operation" OR "co-operations" OR "cooperation" OR "cooperations" OR "Accepting" OR "acceptance" OR "Behavior Control" OR "Behavior Therapy" OR "Problem Behavior" OR "Cooperative Behavior") AND ("parents" OR "parent" OR "Parent-Child Relations" OR "parental" OR "mothers" OR "mother" OR "fathers" OR "father") AND ("child" OR "children" OR "childhood" OR "child, preschool" OR "preschool" OR "preschools" OR "pediatrics" OR "pediatric" OR "paediatrics" OR "paediatric" OR "Child Behavior") AND ((("dental" OR "dentistry") AND ("visit" OR "visits" OR "treatment" OR "treatments" OR "restoration" OR "restorations" OR "Tooth Extraction" OR "Extraction" OR "Extractions" OR "Dental Prophylaxis" OR "Prophylaxis")) OR "Dental Care" OR "Dental Care for Children" OR "Dental Offices" OR "Dental Office" OR "Pediatric Dentistry" OR "oral health") |
| **ProQuest** | noft(("Conditioning, Operant" OR "Operant Conditioning" OR "Operant Conditionings" OR "Instrumental Learning" OR "Restraint, Physical" OR "Physical Restraint" OR "Physical Restraints" OR "Physical Immobilization" OR "Immobilization" OR "Persuasive Communication" OR "Conscious Sedation" OR "Reinforcement(Psychology)" OR "Reinforcement" OR "Reinforcements" OR "Collaboration" OR "Collaborations" OR "co-operation" OR "co-operations" OR "cooperation" OR "cooperations" OR "Accepting" OR "acceptance" OR "Behavior Control" OR "Behavior Therapy" OR "Problem Behavior" OR "Cooperative Behavior") AND ("parents" OR "parent" OR "Parent-Child Relations" OR "parental" OR "mothers" OR "mother" OR "fathers" OR "father") AND ("child" OR "children" OR "childhood" OR "child, preschool" OR "preschool" OR "preschools" OR "pediatrics" OR "pediatric" OR "paediatrics" OR "paediatric" OR "Child Behavior") AND ((("dental" OR "dentistry") AND ("visit" OR "visits" OR "treatment" OR "treatments" OR "restoration" OR "restorations" OR "Tooth Extraction" OR "Extraction" OR "Extractions" OR "Dental Prophylaxis" OR "Prophylaxis")) OR "Dental Care" OR "Dental Care for Children" OR "Dental Offices" OR "Dental Office" OR "Pediatric Dentistry" OR "oral health")) |

**Appendix 2** - Excluded articles and reasons for exclusion (n=32).

| **Author, Year** | **Reason for exclusion** |
| --- | --- |
| Abushal, Adenubi 2009^1^ | 1 |
| Almarwan et al 2018^2^ | 2 |
| Araujo et al 2010^3^ | 1 |
| Arch et al 2001^4^ | 2 |
| Ashley et al 2010^5^ | 2 |
| Bayardo et al 2012^6^ | 1 |
| Blain, Hill 1998^7^ | 4 |
| Chang et al 2018 ^8^ | 5 |
| Chiaretti 2010^9^ | 1 |
| Cohenour et al 1978^10^ | 2 |
| Desai et al^11^ | 4 |
| Elango 2012^12^ | 5 |
| Gomes 2017^13^ | 3 |
| Grewal 2003^14^ | 2 |
| Heinrich 2004^15^ | 2 |
| Jain 2013^16^ | 2 |
| Kaygisiz, Yesil 2000^17^ | 2 |
| Kupietzky 2005^18^ | 5 |
| Lahoud 2001^19^ | 2 |
| Lee et al 2002^20^ | 2 |
| Meira 2009^21^ | 1 |
| Peretz 2014^22^ | 2 |
| Quinby 2004^23^ | 1 |
| Ram et al 2010^24^ | 1 |
| Rodrigues et al^25^ | 1 |
| Shaw et al 1996^26^ | 1 |
| Shroff et al 2015^27^ | 1 |
| Soldani et al 2010^28^ | 2 |
| Veerkamp et al^29^ | 2 |
| White et al 2003^30^ | 1 |
| White et al 2016^31^ | 1 |
| Wood 2010^32^ | 2 |

1) Studies that did not evaluate the parents’ agreement of behavior guidance techniques but instead addressed parents’ satisfaction/preferences and/or success rate and treatment costs;

2) Lacked data regarding parents’ agreement with behavior guidance techniques;

3) Secondary studies (review articles, letters to the editor, books, book chapters etc.);

4) Did not find complete data in published article;

5) Articles that duplicated participants from other publications.

1.	Abushal M, Adenubi JO. Attitudes of Saudi parents toward separation from their children during dental treatment. *The Saudi dental journal*. 2009;21(2):63-67.

2.	Almarwan M. Parental Perception toward Dental Sedation in Pediatric Patients at the University of Maryland [10817539]. Ann Arbor: University of Maryland, Baltimore; 2018.

3.	Araújo SM, Silveira EG, Mello LD, Caregnato M, Dal, Asta VG. Ponto de vista dos pais em relação a sua presença durante o atendimento odontológico de seus filhos. *Salusvita*. 2010;29(2):17-27.

4.	Arch L, Humphris G, Lee G. Children choosing between general anaesthesia or inhalation sedation for dental extractions: the effect on dental anxiety. *International journal of paediatric dentistry*2001. p. 41-48.

5.	Ashley PF, Parry J, Parekh S, Al-Chihabi M, Ryan D. Sedation for dental treatment of children in the primary care sector (UK). *British dental journal*. 2010;208(11):E21; discussion 522-523.

6.	Bayardo RA, Herrera ML, Aceves L. Midazolam conscious sedation in 2-4 years old children. *RGO*. 2012;60(3).

7.	Blain K, Hill F. The use of inhalation sedation and local anaesthesia as an alternative to general anaesthesia for dental extractions in children. *British dental journal*1998.

17.	Kaygisiz YY. Parental understanding and acceptance of behavior management for child dental patients [1397833]. Ann Arbor: University of Minnesota; 2000.

25.	Rodrigues VBM, Costa LR, Corrêa de Faria P. Parents' satisfaction with paediatric dental treatment under sedation: A cross-sectional study. *International journal of paediatric dentistry*. 2020.

26.	Shaw AJ, Meechan JG, Kilpatrick NM, Welbury RR. The use of inhalation sedation and local anaesthesia instead of general anaesthesia for extractions and minor oral surgery in children: a prospective study. *International journal of paediatric dentistry*. 1996;6(1):7-11.

27.	Shroff S, Hughes C, Mobley C. Attitudes and preferences of parents about being present in the dental operatory. *Pediatric dentistry*. 2015;37(1):51-55.

28.	Soldani F, Manton S, Stirrups D, Cumming C, Foley J. A comparison of inhalation sedation agents in the management of children receiving dental treatment: a randomized, controlled, cross-over pilot trial. *International journal of paediatric dentistry*2010. p. 65-75.

29.	Veerkamp JS, Gruythuysen RJ, van Amerongen WE, Hoogstraten J. Dental treatment of fearful children using nitrous oxide. Part 2: The parent's point of view. *ASDC journal of dentistry for children*.59(2):115-119.

30.	White H, Lee JY, Vann Jr WF. Parental evaluation of quality of life measures following pediatric dental treatment using general anesthesia. *Anesthesia progress*. 2003;50(3):105-110.

31.	White J, Wells M, Arheart KL, Donaldson M, Woods MA. A Questionnaire of Parental Perceptions of Conscious Sedation in Pediatric Dentistry. *Pediatric dentistry*. 2016;38(2):116-121.

32.	Wood MN, Manley MCG, Bezzina N, Hassan R. An audit of the use of intravenous ketamine for paediatric dental conscious sedation. *British dental journal*. 2015;218(10):573-577.

**Appendix 3** - Meta-analysis of parents’ acceptance of each behavior guidance technique in non-special heath care needs children evaluated with Visual Analogic Scale where 100 millimeters is well accepted and zero means not accepted (Comprehensive Meta-Analysis Software - Biostat, Englewood, USA). All meta-analyses used Random effect models.

**A - Distraction**

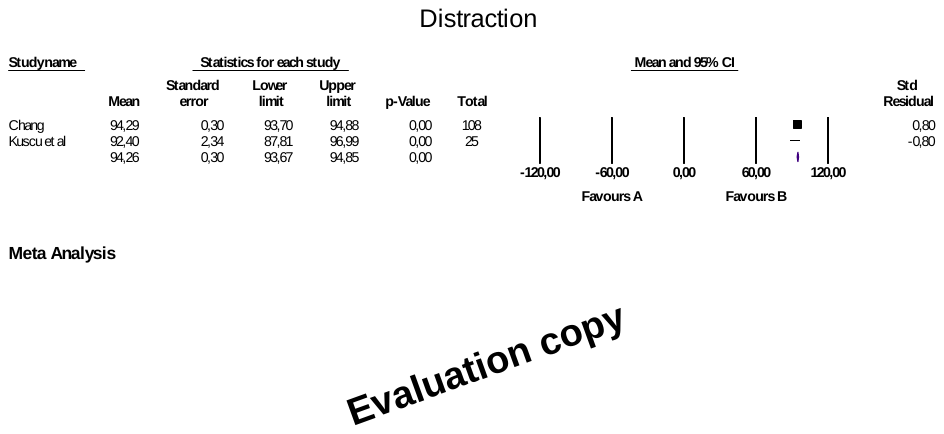

Test for heterogeneity

Q	0.642

DF	1

Significance level	P = 0.423

Inconsistency I2 = 0.00

**B - Positive Reinforcement**

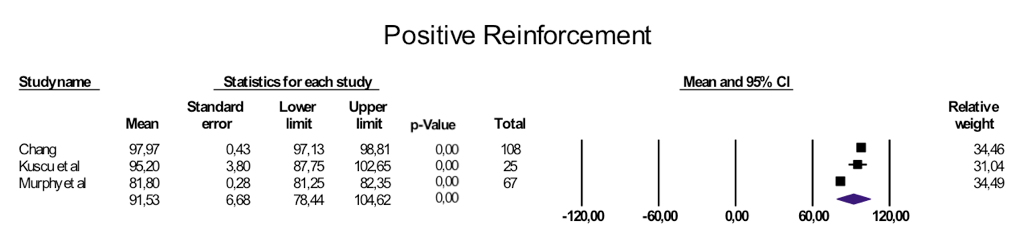

Test for heterogeneity

Q	1001.8

DF	2

Significance level	P = 0.000

Inconsistency I2 = 99.8

**C - Tell-show-do**

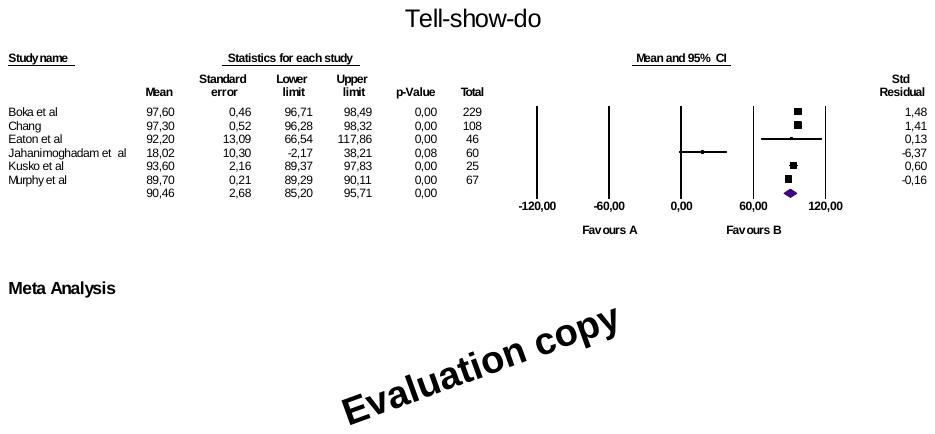

Test for heterogeneity

Q	428.0

DF	5

Significance level	P = 0.000

Inconsistency I2 = 98.8

**D - Nonverbal Communication**

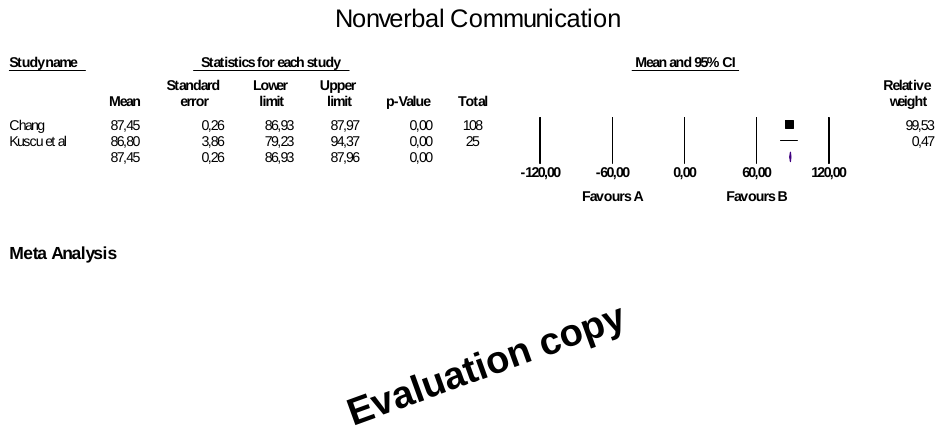

Test for heterogeneity

Q	0.028

DF	1

Significance level	P = 0.86

Inconsistency I2 = 0.00

**E - Nitrous Oxide Inhalation**

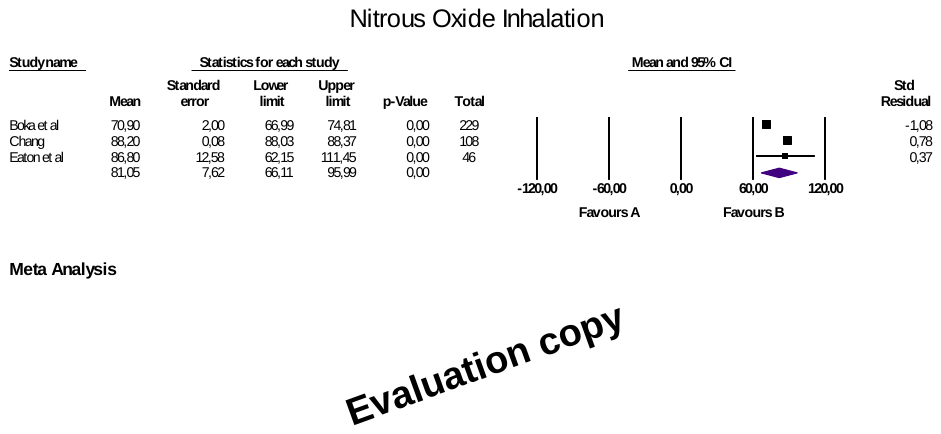

Test for heterogeneity

Q	75.0

DF	2

Significance level	P = 0.00

Inconsistency I2 = 97.3

**F - Parental Presence/Absence**

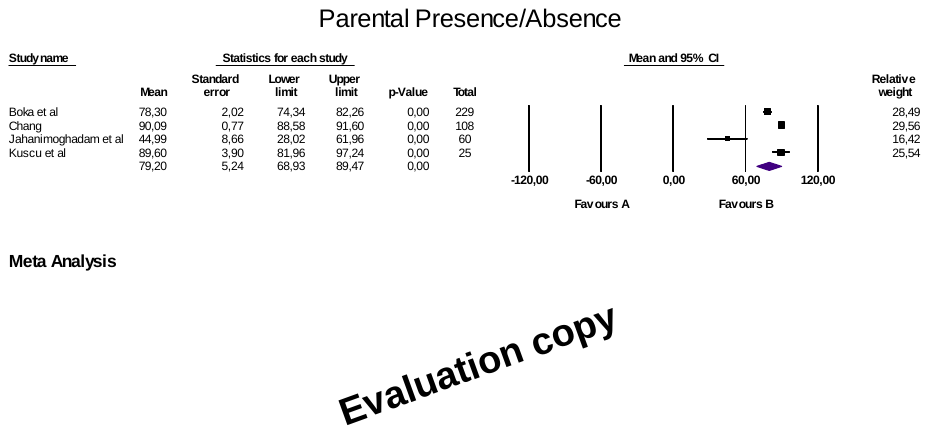

Test for heterogeneity

Q	54.9

DF	3

Significance level	P = 0.00

Inconsistency I2 = 94.5

**G - Voice Control**

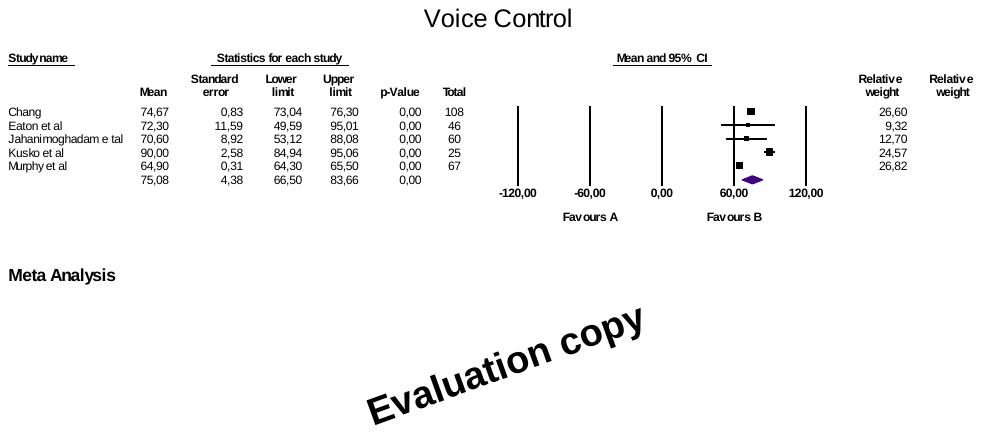

Test for heterogeneity

Q	206.9

DF	4

Significance level	P = 0.00

Inconsistency I2 = 98.0

**H- Sedation**

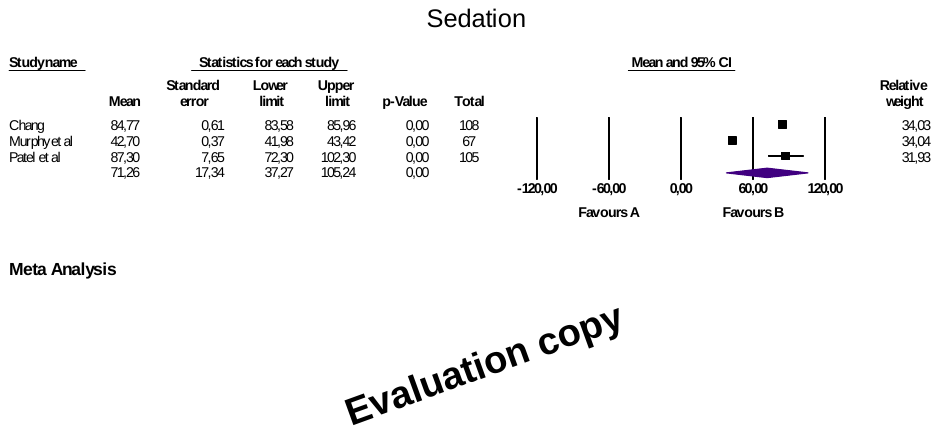

Test for heterogeneity

Q	3529.5

DF	2

Significance level	P = 0.00

Inconsistency I2 = 99.9

**I- Active Protective Stabilization**

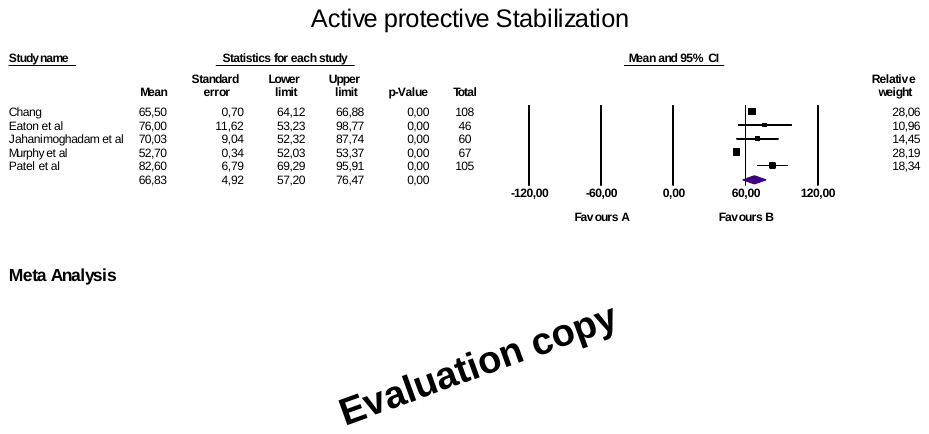

Test for heterogeneity

Q	289.9

DF	4

Significance level	P = 0.00

Inconsistency I2 = 98.6

**J - General Anesthesia**

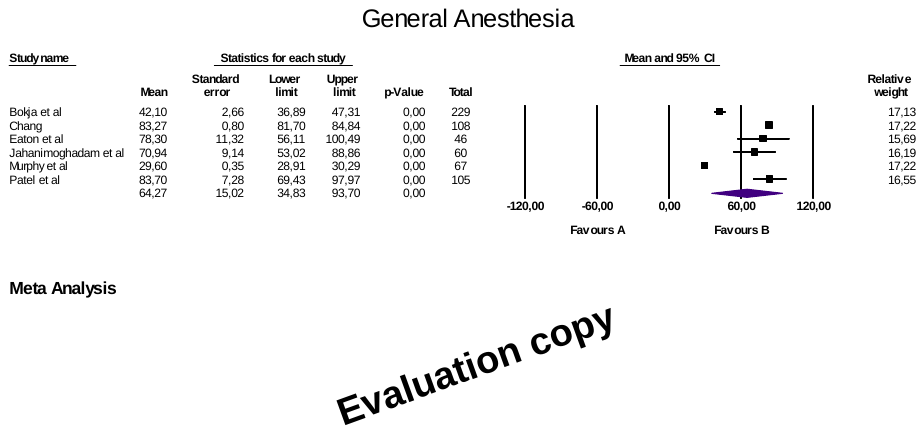

Test for heterogeneity

Q	3823.4

DF	5

Significance level	P = 0.00

Inconsistency I2 = 99.8

**K - Hand-Over-Mouth**

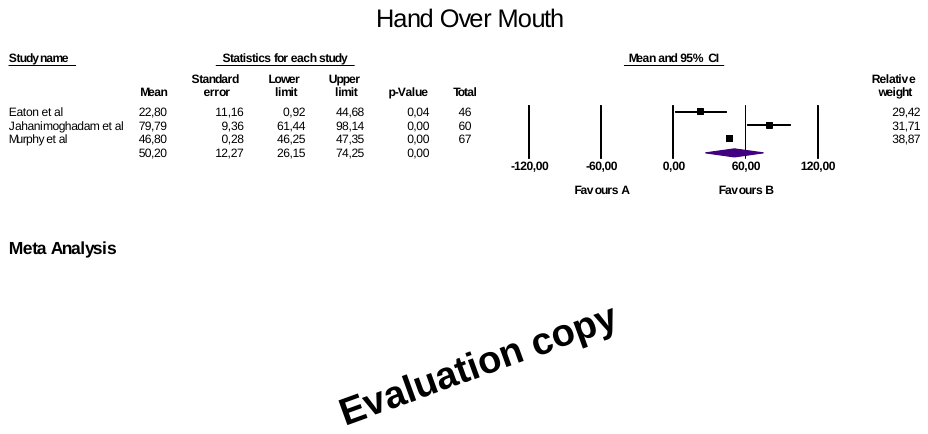

Test for heterogeneity

Q	17.0

DF	2

Significance level	P = 0.00

Inconsistency I2 = 88.2

**L - Passive Protective Stabilization**

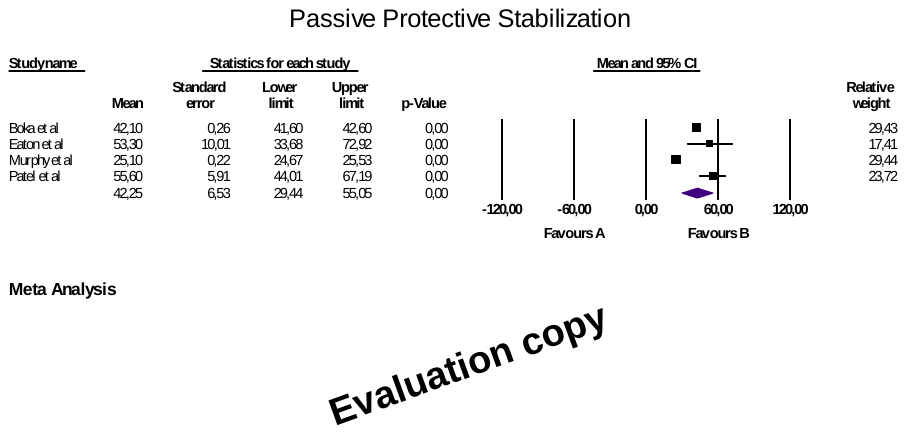

Test for heterogeneity

Q	2560.3

DF	3

Significance level	P = 0.00

Inconsistency I2 = 99.8

**Appendix 4** - Forests plots for the direct comparison of difference in means of acceptance of behavior guidance techniques among parents of non-special health care needs children versus acceptance of parents of special health care needs children measured in millimeters in Visual Analogic Scale. On this scale, zero represents the least acceptable and 100 mm the most acceptable (n=245): A - Hand over the mouth; B - Active protective stabilization; C - Sedation; and D - General anesthesia.

A - Hand over the mouth

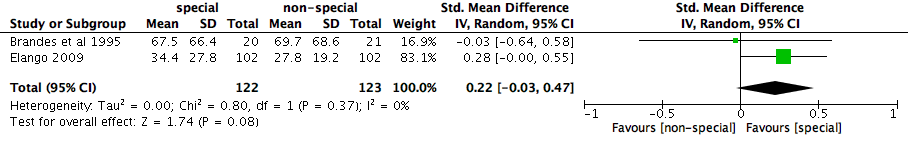

B - Active protective stabilization

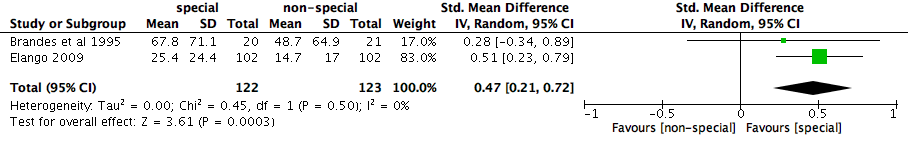

C - Sedation

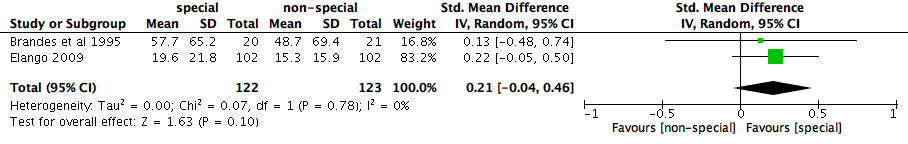

D - General anesthesia

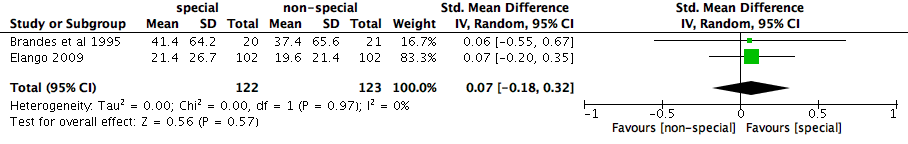

**Appendix 5** - Forests plots for the comparison of acceptance of behavior guidance techniques among parents of non-special health care needs children who received explanation on the techniques versus those who did not receive explanation prior to judge the behavior guidance technique (BGT). Ratings were measured in millimeters on a Visual Analogic Scale where zero represented the most acceptable and 100 mm the least acceptable BGT (n=112): A - Hand over the mouth exercise; B - active protective stabilization; C - Nitrous oxide/oxygen inhalation; D - General anesthesia; E - Passive protective stabilization; F - Oral premedication; G - Voice control; and H - Tell-show-do. Forest plot of difference in means data.

A - Hand over the mouth

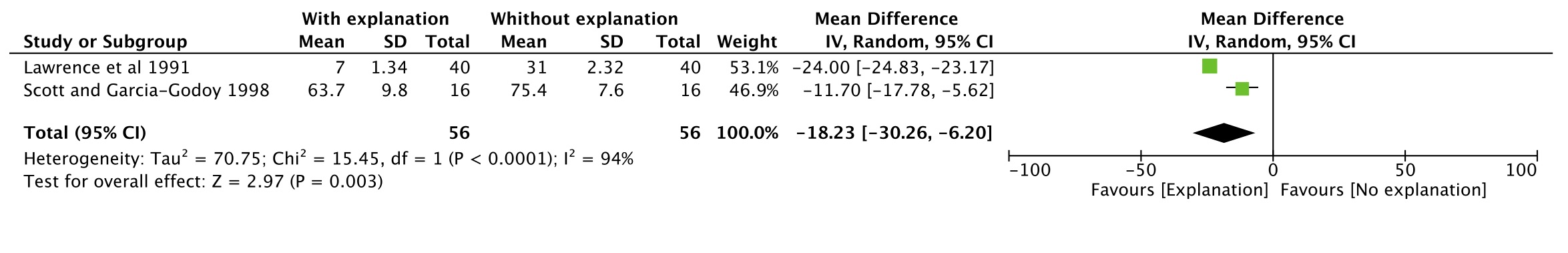

B - Active protective stabilization

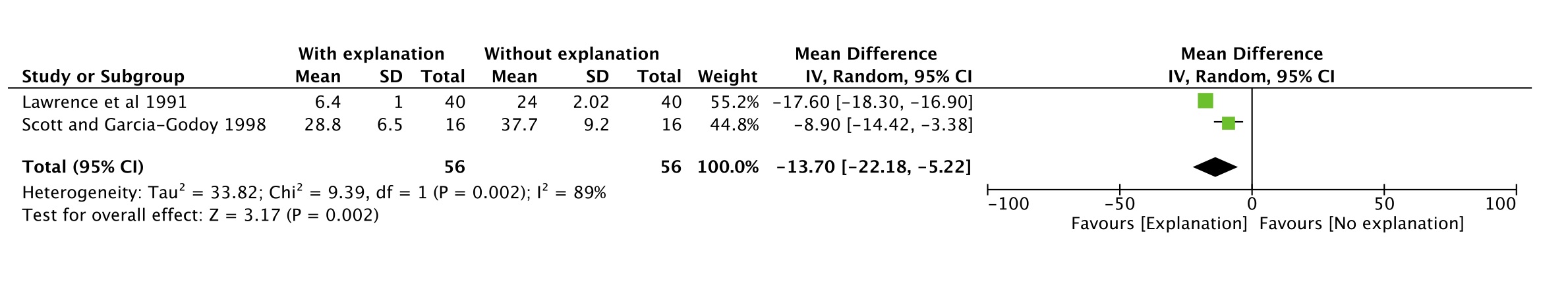

C - Nitrous oxide/oxygen inhalation

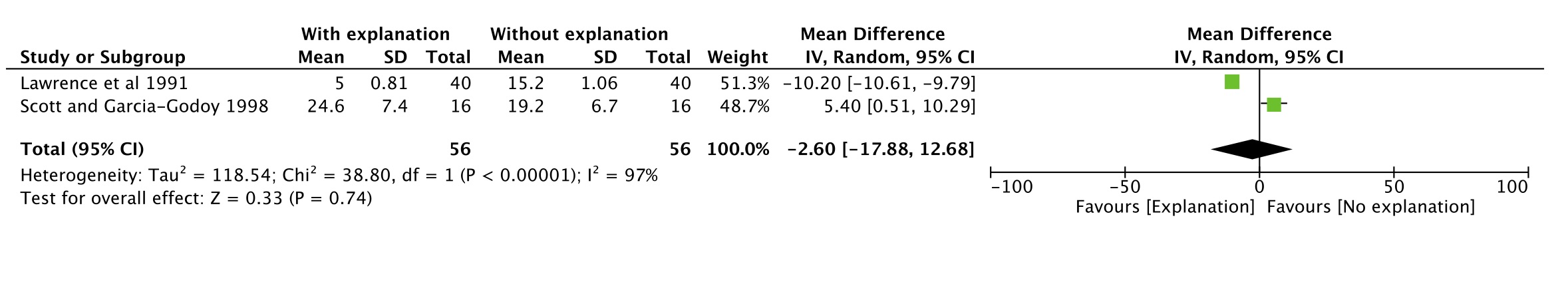

D - General anesthesia

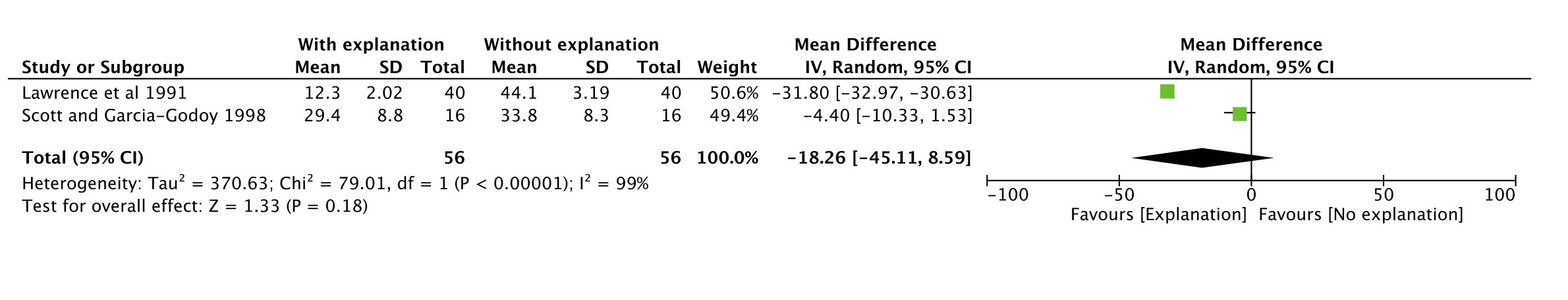

E - Passive protective stabilization

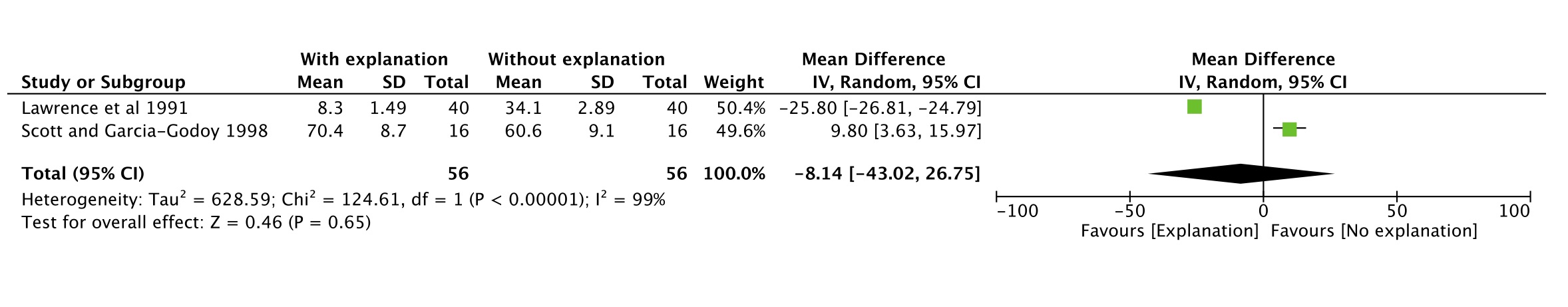

F - Oral premedication

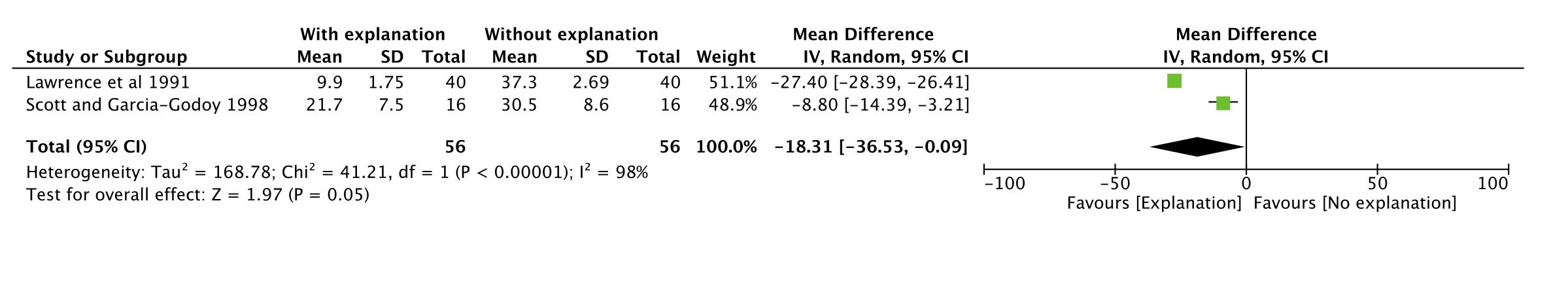

G - Voice control

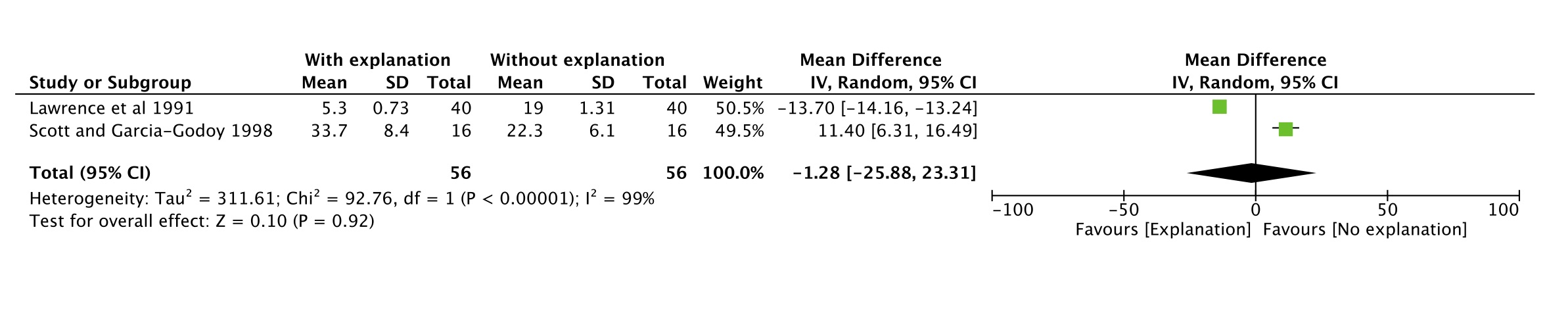

H - Tell-show-do

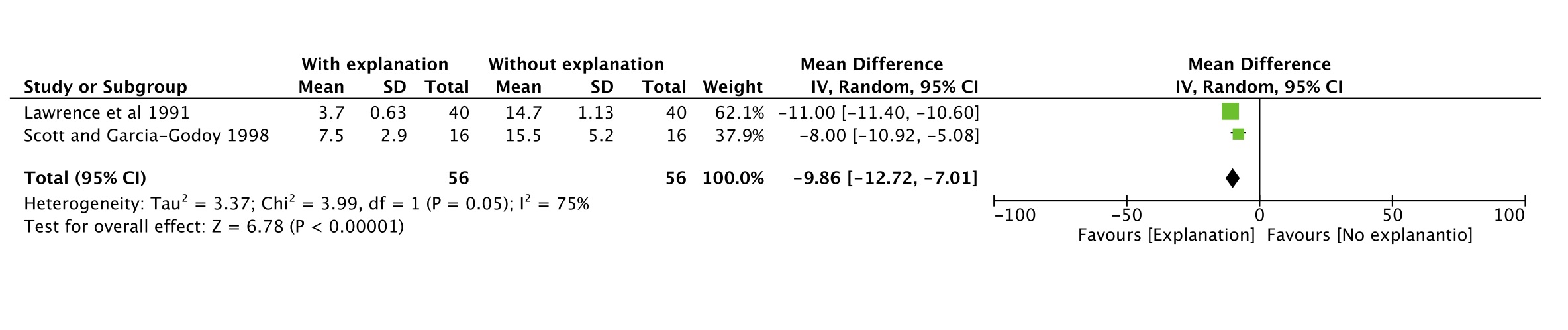

#### Appendix 6. Summary of findings table- GRADE

| **Summary of findings:** | | | |
| --- | --- | --- | --- |
| **Comparison of parental acceptance between children with special health care needs (SHCN) and children without SHCN toward behavior guidance techniques for pediatric dental visits** | | | |
| Outcome № of participants (studies) | |  | Certainty |
| Proportion of non-SHCN children parent’s agreement with BGT for pediatric dental visits. № of participants: (29 observational studies) | Sixteen different behavior guidance technique evaluated in 2594 participants (dichotomous outcome -yes/no) | | ⨁◯◯◯ VERY LOW ^a,b,c^ |
| Proportion of SHCN children parent’s agreement with BGT for pediatric dental visits. № of participants: (5 observational studies) | Nine different behavior guidance technique evaluated in 748 participants (dichotomous outcome -yes/no) | | ⨁◯◯◯ VERY LOW ^a,b,c^ |
| Comparison of acceptance of BGT among parents of non-SHCN and those of SHCN children № of participants: (2 observational studies) | Four different behavior guidance technique evaluated in 245 participants (continuous outcome - means of agreement with BGT) | | ⨁◯◯◯ VERY LOW ^a,d^ |
| Difference in means of agreement with the BGT measured with VAS among parents of non-SHCN who received explanation before the presentation of the technique and those who did not № of participants: (2 observational studies) | Eight different behavior guidance technique evaluated in 112 participants (continuous outcome - means of agreement with BGT) | | ⨁◯◯◯ VERY LOW ^a,d,e^ |
| **GRADE Working Group grades of evidence** **High certainty:** We are very confident that the true effect lies close to that of the estimate of the effect **Moderate certainty:** We are moderately confident in the effect estimate: The true effect is likely to be close to the estimate of the effect, but there is a possibility that it is substantially different **Low certainty:** Our confidence in the effect estimate is limited: The true effect may be substantially different from the estimate of the effect **Very low certainty:** We have very little confidence in the effect estimate: The true effect is likely to be substantially different from the estimate of effect | | | |

#### Explanations

a. Definition of eligibility criteria and confounding factor were missing

b. I2 varied from 32.5 to 98.1%

c. Wide confidence intervals

d. Less than 400 observations for continuous measures

e. I2 above 75%
